# Integrating Causal Discovery and Clinically-Relevant Insights to Explore Directional Relationships between Autistic Features, Sex at Birth, and Cognitive Abilities

**DOI:** 10.1101/2023.12.21.23300348

**Authors:** Sunday Francis, Angela Tseng, Eric Rawls, Christine Conelea, Nicola Grissom, Erich Kummerfeld, Sisi Ma, Suma Jacob

**Author notes:** Correspondence concerning this article should be addressed to: Suma Jacob, M.D., Ph.D., Department of Psychiatry and Behavioral Sciences, University of Minnesota, Minneapolis, MN, 55455, USA.

## Abstract

Prevalence in autism spectrum disorder (ASD) diagnosis has long been strongly male-biased. Yet, consensus has not been reached on mechanisms and clinical features that underlie sex-based discrepancies. Whereas females may be under-diagnosed because of inconsistencies in diagnostic/ascertainment procedures (sex-biased criteria, social camouflaging), diagnosed males may have exhibited more overt behaviors (e.g., hyperactivity, aggression) that prompted clinical evaluation. Applying a novel network-theory-based approach, we extracted data-driven, clinically-relevant insights from a large, well-characterized sample (Simons Simplex Collection) of 2175 autistic males (Ages = 8.9±3.5 years) and 334 autistic females (Ages = 9.2±3.7 years). Exploratory factor analysis (EFA) and expert clinical review reduced data dimensionality to 15 factors of interest. To offset inherent confounds of an imbalanced sample, we identified a subset of males (N=331) matched to females on key variables (Age, IQ) and applied data-driven CDA using Greedy Fast Causal Inference (GFCI) for three groups (All Females, All Males, and Matched Males). Structural equation modeling (SEM) extracted measures of model fit and effect sizes for causal relationships between sex, age, and, IQ on EFA-selected factors capturing phenotypic representations of autism across sensory, social, and restricted and repetitive behavior domains. Our methodology unveiled sex-specific directional relationships to inform developmental outcomes and targeted interventions.

## INTRODUCTION

Autism Spectrum Disorder (ASD) is a complex neurodevelopmental condition characterized by impairments in social communication and interactions, and restricted, repetitive patterns of behavior, interests, or activities (RRBs) which may include atypical interest in sensory-related features of the environment (APA, 2013). While a strong male bias (4.2 male: 1 female ratio) in ASD prevalence has been reported consistently (Maenner et al., 2020), researchers have yet to reach consensus on the mechanisms and clinical features that underlie these sex (at birth) discrepancies (Halladay et al., 2015). Moreover, recent estimates suggest that the veracious male-to-female ratio of ASD may be nearer to 2– 3: 1 (Lai, Lombardo, Auyeung, Chakrabarti, & Baron-Cohen, 2015; Posserud, Skretting Solberg, Engeland, Haavik, & Klungsøyr, 2021), attributed, in part, to under-diagnosis or delayed recognition of autism in girls and women overall (Loomes, Hull, & Mandy, 2017; Whitlock, Fulton, Lai, Pellicano, & Mandy, 2020) and impacting their timely access to services and supports (Bargiela, Steward, & Mandy, 2016). Lifespan estimates (1.8 male: 1 female) have been reported (Rutherford et al., 2016) reflecting significant differences in the mean age of referral and diagnosis for girls compared to boys, add further evidence of delayed recognition of ASD in girls.

Given that diagnostic criteria for ASD have been informed historically by male models of autism, prevailing assessments focusing on paradigmatic autistic features may fail to identify more nuanced female presentations (Stephenson, Norris, & Butter, 2023; Wood-Downie, Wong, Kovshoff, Cortese, & Hadwin, 2021). For example, whereas autistic females are more likely to experience internalizing symptoms (e.g., anxiety, depression), autistic males often exhibit more disruptive externalizing behaviors (e.g., hyperactivity, aggression) that prompt earlier clinical evaluations and diagnoses (Lai et al., 2015; Mandy et al., 2011). Sociocultural factors such as higher expectations for females to engage in social communication and interactions may also bias ascertainment of autistic features (Kreiser & White, 2013). Females are more likely to demonstrate ‘camouflaging’ behaviors during human interactions such that they may to employ strategies to hide their autistic characteristics to fit into ‘neurotypical’ social environments (Ai, Cunningham, & Lai, 2022; Hull, Petrides, et al., 2017; Livingston, Colvert, Bolton, & Happé, 2019). Unfortunately, although camouflaging may be initially adaptive for social adjustment, later diagnosis, along with the long-term stress of effortful masking and compensation, have been associated with negative mental health and increased risk for suicidality (Cassidy, Bradley, Shaw, & Baron-Cohen, 2018; Hull, Petrides, & Mandy, 2020). Hence, it is critical that we improve our understanding and recognition of sex-based phenotypic profiles in autism to provide targeted support for males and females at critical early developmental stages (Lai & Szatmari, 2020).

While the prevalence of ASD in females may be closer to males than previously estimated, phenotypically differing sex/gender^1^ profiles in ASD have been observed at an early age. Historically, more intellectual, emotional, and behavioral challenges have been reported in autistic females than males (Duvekot et al., 2017; Frazier, Georgiades, Bishop, & Hardan, 2014; Russell et al., 2022), and these observations have contributed to the specious syllogism that autistic females are more likely to show neurological and functional impairments than males (de Giambattista, Ventura, Trerotoli, Margari, & Margari, 2021; Kaat et al., 2020). Yet, the alternate view indicates that females need to demonstrate more severe developmental, behavioral, or intellectual disability to be diagnosed with ASD because the female phenotype manifests in a more oblique fashion. For example, children with better communication abilities are diagnosed with autism significantly later than non-verbal and minimally verbal children; girls with complex phrase speech are also diagnosed later than boys with comparable verbal skill levels (Salomone, Charman, McConachie, & Warreyn, 2016), potentially because higher functioning girls utilize camouflaging strategies to appear less functionally impaired. Consequently, more profound concerns may have to be expressed before females are referred for clinical evaluations.

Although large scale studies have reported a great deal of variation in social communication and cognitive abilities across sexes/genders in ASD (Hull, Mandy, & Petrides, 2017; Tillmann et al., 2018), observations of sex-specific presentations of RRBs have been persistent. RRBs represent a heterogeneous cluster of behavioral symptoms that include restricted interests, preoccupation with parts of objects, repetitive motor mannerisms, insistence on sameness, sensory behaviors, and strict adherence to specific routines or rituals. Relative to the well-studied domain of social communication, less is known about how RRBs vary according to individual characteristics, including sex and cognitive ability (Frazier et al., 2014; Hartley & Sikora, 2009; Van Wijngaarden-Cremers et al., 2014). As supported by the extant literature broadly, males present with higher levels of RRBs than females (Supekar & Menon, 2015; Szatmari et al., 2012). Given that ASD samples have been predominantly male, a distinct “female” expression (i.e., with less emphasis on RRBs) of the underlying biological liability in autism may be overlooked clinically.

Increasing support for discernable sex-based expressions of autistic characteristics, particularly in the domain of RRBs, has highlighted a need for improved recognition and understanding of mechanisms underlying sex/gender discrepancies in ASD phenotypic profiles. Given the diverse symptomatology, a data-driven approach is critical to disentangle the complex interplay between biological, psychological, and environmental mechanisms underlying male and female presentations in autism (Maxwell, Harrison, Rawls, & Zilverstand, 2022). Novel implementations of network theory have shown promise in constructing and analyzing causal or directional relations between symptoms in psychopathology, factoring in strength of interactions as an additional endophenotype (Borsboom, 2017; Borsboom & Cramer, 2013). Accordingly, to investigate the potential of a network approach in ASD, we applied exploratory factor analysis (EFA) and causal discovery analysis (CDA) to model the structure and relationships between factors subserving core autism features in a large, well-characterized ASD dataset (Simons Simplex Collection v15.3; SSC). Considering the broad expanse of the autistic symptom space and the relative paucity of research in the RRB domain (compared to social communication), we reduced our data dimensionality by focusing on RRB factors and integrating clinically-relevant insights into our analyses. Taken together, our approach leveraging clinician expertise with CDA affords us an efficient means to infer meaningful causal connections from statistical relationships in data (Spirtes, Glymour, & Scheines, 2000).

## METHODS

Our behavioral dataset of well-characterized autistic youth was obtained from the SSC v15.3 database; data collection and methodology has been described in detail in previously published reports (Fischbach & Lord, 2010) and the website; http://sfari.org/simons-simplex-collection). Participation in the SSC included diagnostic evaluation, collection of phenotypic measures, and cognitive assessment. Data collection, entry, and validation methods were standardized across collection sites to ensure data reliability. Informed consent was obtained during the original data collection stage and participants opted in to include their de-identified data in further investigations. Analyses included all participants who completed our measures of interest with the total sample consisting of 2509 individuals, predominantly male (86.7%), aged 4-18 years old with ASD. With the goal of detecting interpretable differences in sex-specific presentations, we designated three subgroups for comparison: 1) all females (F: N = 334; Age = 9.2 ± 3.7 years), 2) all males (AM: N = 2175; Age = 8.9 ± 3.5 years), and 3) and matched males (MM: N = 331; Age = 8.1 ± 3.2 years), a subset of males exact-matched individually by full scale IQ (FSIQ) and by nearest-match to AGE, verbal IQ (VIQ), and non-verbal IQ (NVIQ) to all females in our dataset. We used the R *matchit* function (Ho, Imai, King, & Stuart, 2011) to define the matched male subgroup. Multiple variable combinations were gauged for best approach; distance in the matching method was estimated using logistic regression. (See SM for additional matching details). Table 1 shows participant subgroup characteristics.

**Table 1.**
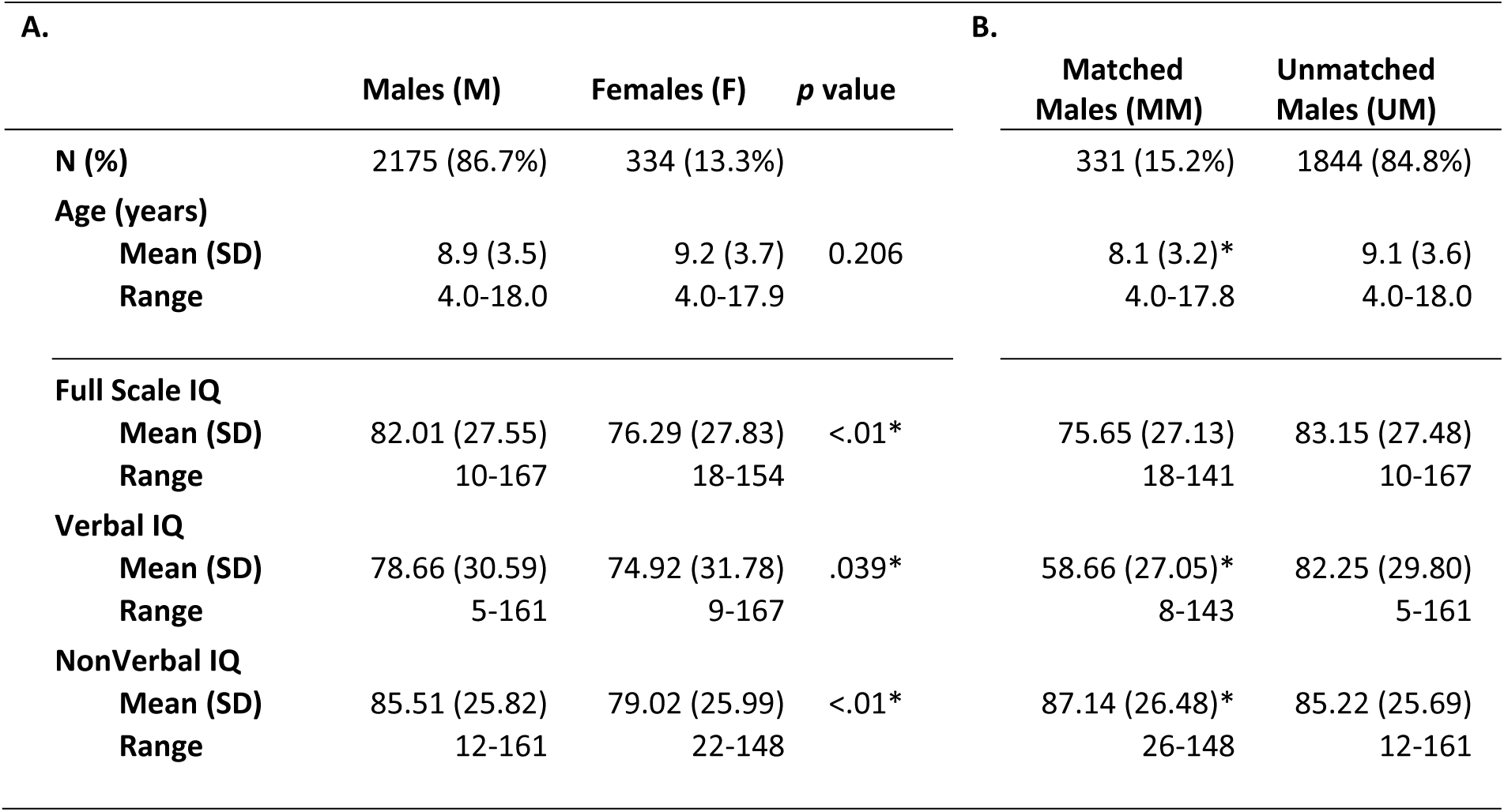
Demographic characteristics of: **(A.)** all Males (M) and all Females (F) in total sample, *p v*alue for M vs. F comparison; **(B.)** subset of males matched to females (Matched Males) and all remaining males (Unmatched Males). (* indicates significant difference (*p* < 0.05) from all Females)

| **A.** |  |  |  | **B.** |  |
| --- | --- | --- | --- | --- | --- |
|  | **Males (M)** | **Females (F)** | **$p$ value** | **Matched Males (MM)** | **Unmatched Males (UM)** |
| **N (%)** | 2175 (86.7%) | 334 (13.3%) |  | 331 (15.2%) | 1844 (84.8%) |
| **Age (years)** |  |  |  |  |  |
| **Mean (SD)** | 8.9 (3.5) | 9.2 (3.7) | 0.206 | 8.1 (3.2)* | 9.1 (3.6) |
| **Range** | 4.0-18.0 | 4.0-17.9 |  | 4.0-17.8 | 4.0-18.0 |
| **Full Scale IQ** |  |  |  |  |  |
| **Mean (SD)** | 82.01 (27.55) | 76.29 (27.83) | <.01* | 75.65 (27.13) | 83.15 (27.48) |
| **Range** | 10-167 | 18-154 |  | 18-141 | 10-167 |
| **Verbal IQ** |  |  |  |  |  |
| **Mean (SD)** | 78.66 (30.59) | 74.92 (31.78) | .039* | 58.66 (27.05)* | 82.25 (29.80) |
| **Range** | 5-161 | 9-167 |  | 8-143 | 5-161 |
| **NonVerbal IQ** |  |  |  |  |  |
| **Mean (SD)** | 85.51 (25.82) | 79.02 (25.99) | <.01* | 87.14 (26.48)* | 85.22 (25.69) |
| **Range** | 12-161 | 22-148 |  | 26-148 | 12-161 |

### Behavioral Measures

To ensure sufficient observations of RRBs in our dataset, we compiled data collected from the SSC cohort using three parent-report assessments that address RRBs directly: the Aberrant Behavior Checklist-Community Version (ABC-CV), the Repetitive Behavior Scale-Revised (RBS-R), and the Social Responsiveness Scale (SRS). The ABC-CV (Aman et al., 1985) is an empirically developed, five-factor, 53-item questionnaire that assesses symptoms of irritability and agitation, social withdrawal or lethargy, stereotypic behavior, hyperactivity and non-compliance, and inappropriate speech. The RBS-R (Bodfish et al, 1995, 2000; Cuccaro et al, 2003, 2007; Miranda et al, 2010) determines the presence of, characterization, and severity of RRBs in individuals with neurodevelopmental disorders (NDDs). The 43-item Likert-scale questionnaire yields six subscales from a completed form - stereotypy, self-injurious behavior (SIB), compulsive behaviors, ritualistic behaviors, insistence on sameness, and restricted behaviors. Parents also completed the SRS (Constantino & Gruber, 2005), a 65-item, Likert-scale questionnaire, measuring reciprocal social behavior. The completed assessment yields a total score and five subscale scores (social awareness, social cognition, social communication, social motivation, and RRBs). Greater severity of social deficits is demonstrated by higher scores. Each item from each questionnaire was included in our EFA.

### Exploratory Factor Analysis (EFA)

Using the ‘psych’ package for R (Revelle, 2017), we reduced data dimensionality from all included item-level responses (161 items total) by applying EFA with maximum likelihood factor extraction and direct oblimin rotation (to allow for correlated factors). Monte Carlo permutation analysis (parallel analysis) was implemented to determine the number of factors to retain (Horn, 1965); using 1000 permutations of the raw data, we retained factors with eigenvalues greater than the 95th percentile of permutation eigenvalues (Glorfeld, 2016). The 23 factors that met our criterion (Supplementary Figure 1) were further reduced by ordering each loading within a factor greatest to smallest. Only items with loadings = |x|< 0.30 were considered as contributing to the factor. Lower variance factors were removed if an item also appeared in a higher variance factor (a full list of factors and items are located in Supplement Table 1) These steps yielded 15 final factors for continued analyses (see Table 2).

**Table 2.** Exploratory Factor Analysis (EFA) and Clinical Consensus Derived Factors of Interest

| Factor # | Label Name | 1. Assessments | Domains |
| --- | --- | --- | --- |
| F-01 | Oppositional (Outburst Behaviors) | ABC | SCI |
| F-02 | Isolated (Alone Preferred) | ABC;SRS | SCI |
| F-03 | Hand/Body Movements (Recurring Mannerisms, Stereotypies) | ABC;RBS-R;SRS | RRBs |
| F-04 | Self-Injurious Behaviors | ABC;RBS-R | RRBs, Sens |
| F-05 | Inflexible (Insistent Behaviors) | RBS-R;SRS | RRBs |
| F-06 | Social Atypicalities (Awkward, Odd Responses) | SRS | SCI |
| F-07 | Motor Overflow (Excessive Impulsive Activity) | ABC;RBS-R | RRBs |
| F-08 | Repetitive Speech (Perseverative Vocal Overflow) | ABC;RBS-R;SRS | RRBs, SCI, Sens |
| F-10 | Staring (into Space; Preoccupied) | ABC;SRS | Sens |
| F-12 | Socioemotional Unresponsiveness | ABC;SRS | SCI, Sens |
| F-14 | Body/Head Movements (Repetitive Rocking/Turning) | ABC;RBS-R | RRBs |
| F-15 | Socioemotional Awareness (Responsive/Expressive) | SRS | RRBs |
| F-16 | Self-Confidence (Social Communication & Interaction) | SRS | SCI |
| F-17 | Sensory & Object Preoccupation | SRS;RBS-R | Sens |
| F-23 | Obsessive Compulsive Behaviors | RBS-R | RRBs, Sens |

Assessments: Aberrant Behavior Checklist-Community Version (ABC-CV); Repetitive Behavior Scale-Revised (RBS-R); Social Responsiveness Scale (SRS).
Domains: Social Communication & Interaction (SCI); Restricted, Repetitive Patterns of Behavior, Interests, or Activities (RRBs); Sensory Sensitivity (Sens)

Two expert clinicians reviewed all items independently and assigned factors into clinically relevant domains of social communication and interaction (SCI), RRBs, and sensory sensitivity (Sens). Consensus reviews were conducted; inter-rater reliability for category matching was > 0.8. For a few factors (see Table 2 and Table S1), reviewers concurred that certain items from the sensory domain overlapped with items from the RRB domain (obsessive compulsive behaviors or self-injurious behaviors), with items from SCI (socioemotional unresponsiveness), or with items from both RRB and SCI (repetitive speech).

### Causal Discovery Analysis: Greedy Fast Causal Inference

We used the Greedy Fast Causal Inference (GFCI; freely available Java program Tetrad 6.7.0 (https://github.com/cmu-phil/tetrad) algorithm (Ogarrio, Spirtes, & Ramsey, 2016) to identify the causal structure (qualitative causal relationship, e.g., A is a direct cause of B) that best fits the data. GFCI consists of a two-step process that searches the space of possible causal structures underlying observed relationships between variables. In addition to determining the set of all probabilistic causal relationships among a set of input variables, GFCI can also detect and depict the potential presence of underlying (unmeasured) confounding factors that may explain putatively causal relationships in complex data. In the first step, GFCI searches potential causal models, applying a fast score-based algorithm (Fast Greedy Equivalence Search; FGES) to assign a likelihood score (Bayesian Information Criterion; BIC) that penalizes overly complex models (Chickering, 2002; Ramsey, 2015). During the FGES phase, GFCI greedily adds then removes edges to maximize the model fit. Second, GFCI performs a series of conditional independence tests to rule out preliminary causal relationships not borne out by the data. Prior to analysis, we restricted our model by removing impossible causal relationships, e.g., sex causing age, IQ causing sex, or age causing sex. Model restriction was achieved via the use of tiered knowledge in Tetrad. GFCI parameters were set using a “penalty discount” of 1 to compute penalized likelihoods in the first step (corresponding to the standard Bayesian Information Criteria; BIC) (Schwarz, 1978) and a Fisher *Z p*-value of 0.01 to conduct conditional independence tests in the second step. These are the default settings for these parameters in Tetrad and are typical parameters for applied data analysis.

Using a combination of goodness of fit statistics (BIC) and conditional independence tests (Fisher’s Z), GFCI analyses identify the best fitting models of a causal process, including the possibility of latent common causes (Spirtes et al., 2000), and represents these output visually as Partial Ancestral Graphs (PAGs). In PAGS, variables are represented as nodes while the type and orientation of connections between two nodes specify the nature of modeled causal relationships (Chickering, 2002; Ogarrio et al., 2016; Ramsey, 2015; Spirtes et al., 2000).

### Effect Size Estimation and Model Fit Statistics

To recover effect sizes for causal relationships (e.g., the direct effect of A on B is the amount of change in variable B when variable A is changed by 1 unit while other variables are held constant), we built a Structural Equation Model (SEM) of the GFCI results using the ‘lavaan’ package (Rosseel, 2012) for R (Ramsey, 2015). Raw and standardized effect sizes were estimated by fitting a linear SEM to the PAG. Causal relationships detected by GFCI were included as direct paths in the SEM while confounded or otherwise uncertain relationships were included as covariances.

Graph stability was assessed from 1,000 bootstrap samples, applying the same GFCI analysis to each, and aggregating the resulting 1,000 graphs into a table summarizing the proportion of all possible relationships (See Table S2). Edges (i.e., connections between variables) were classified as directed (in either direction), semi-directed (in either direction), undirected, or bi-directed. Bootstrap values indicating the proportion of each edge presence in resampling represent stability or consistency of each connection (moderate ≥ 50%; high ≥ 75%). The highest frequency edge type by each variable pair was identified by ensemble rule (Soltis & Soltis, 2003; Stevenson, Kummerfeld, & Merrill, 2021). Edges with an absolute estimated effect size of at least 0.1 were retained to direct focus on relationships with meaningful strength without overlooking potential (but weaker) connections (strong: *r* ≥ 0.50; moderate: *r* ≥ 0.30) (Anker, Kummerfeld, Rix, Burwell, & Kushner, 2019; Ogarrio et al., 2016; Stevenson et al., 2021).

### Sample Subgroups

Separately for Females (F), Matched Males (MM), and All Males (AM), we built data-driven causal models of relationships between factors, both predetermined (age, verbal IQ; VIQ, non-verbal IQ; NVIQ) and discovered, that underlie ASD-associated behaviors. We then compared the causal models generated for each subgroup to examine sex differences in patterns of ASD symptomatology. Separate graphs for each subgroup were qualitatively compared for edges that were present in one model but not in the other, or edges that were present in more than one model but with different orientations. Edge stability and SEM effect sizes were used to quantitatively compare the strength and direction of each causal relationship.

## RESULTS

To investigate the role of sex (at birth) on causal relationships between AGE, NVIQ, and VIQ and EFA-derived factors across the SCI, RRB, and Sens domains, we compared PAGs for Female Only, Matched Male, and All Male subgroups, focusing on common pathways across sub-groups. We report bootstrap values (stability/consistency) and standardized edge weights (effect size) of each relationship; *p* values were uniformly <0.001 for all edges due to large sample size and preference for CDA to generate sparse models with strong pathways.

### ALL GROUPS: FEMALES & ALL MALES & MATCHED MALES (COMMON PATHS)

Several pathways (presence and direction) were consistent across all groups or directionally across the entire sample. For example, increasing AGE related to a decrease in Motor Overflow (Excessive Impulsive Activity) (F: 0.90, -0.39; AM: 0.38, -0.27; MM: 0.58, -0.22). Increased NVIQ associated with reduced Hand/Body Movements (Recurring Mannerisms, Stereotypies) (F: 0.72, -0.35; AM: 0.37, -0.32; MM: 0.45, -0.26) and Sensory & Object Preoccupation (F: 0.72, -0.37; AM: 0.88, -0.25; MM: 0.95, -0.40); higher NVIQ was linked to higher Self-Confidence (SCI) (F: 0.49, 0.23; AM: 0.93, 0.17; MM: 0.22, 0.28). Being more Isolated (Alone Preferred) related to increased Socioemotional Unresponsiveness (F: 0.60, 0.38; AM: 0.72, 0.31; MM: 0.72, 0.39) and increased Socioemotional Awareness (Responsive/Expressive) related to a decrease in Social Atypicalities (Awkward, Odd Responses) (F: 0.75, -0.34; AM: 0.47, -0.27; MM: 0.34, -0.26). Further, more Body/Head Movements (Repetitive Rocking/Turning) were associated with more Motor Overflow (Excessive Impulsive Activity) (F: 0.46, 0.26; AM: 0.56, 0.17; MM: 0.35, 0.29) and more Hand/Body Movements (Recurring Mannerisms, Stereotypies associated with an increase in Body/Head Movements (Repetitive Rocking/Turning) (F: 0.71, 0.56; AM: 0.65, 0.49; MM: 0.63, 0.52).

### FEMALES & MATCHED MALES (COMMON PATHS)

Common pathways (Figure 1) were found between the female and matched male subgroups, such that increased AGE related to a decrease in Motor Overflow (Excessive Impulsive Activity) (F: 0.90, - 0.39; MM: 0.58, -0.22) as well as an increase in Social Atypicalities (Awkward, Odd Responses) (F: 0.89, 0.37; MM: 0.79, 0.28). Higher NVIQ was associated with a decrease in Hand/Body Movements (Recurring Mannerisms, Stereotypies) (F: 0.72, -0.35; MM: 0.45, -0.26) and decreased Sensory & Object Preoccupation (F: 0.72, -0.37; MM: 0.95, -0.40).

**Figure 1.**
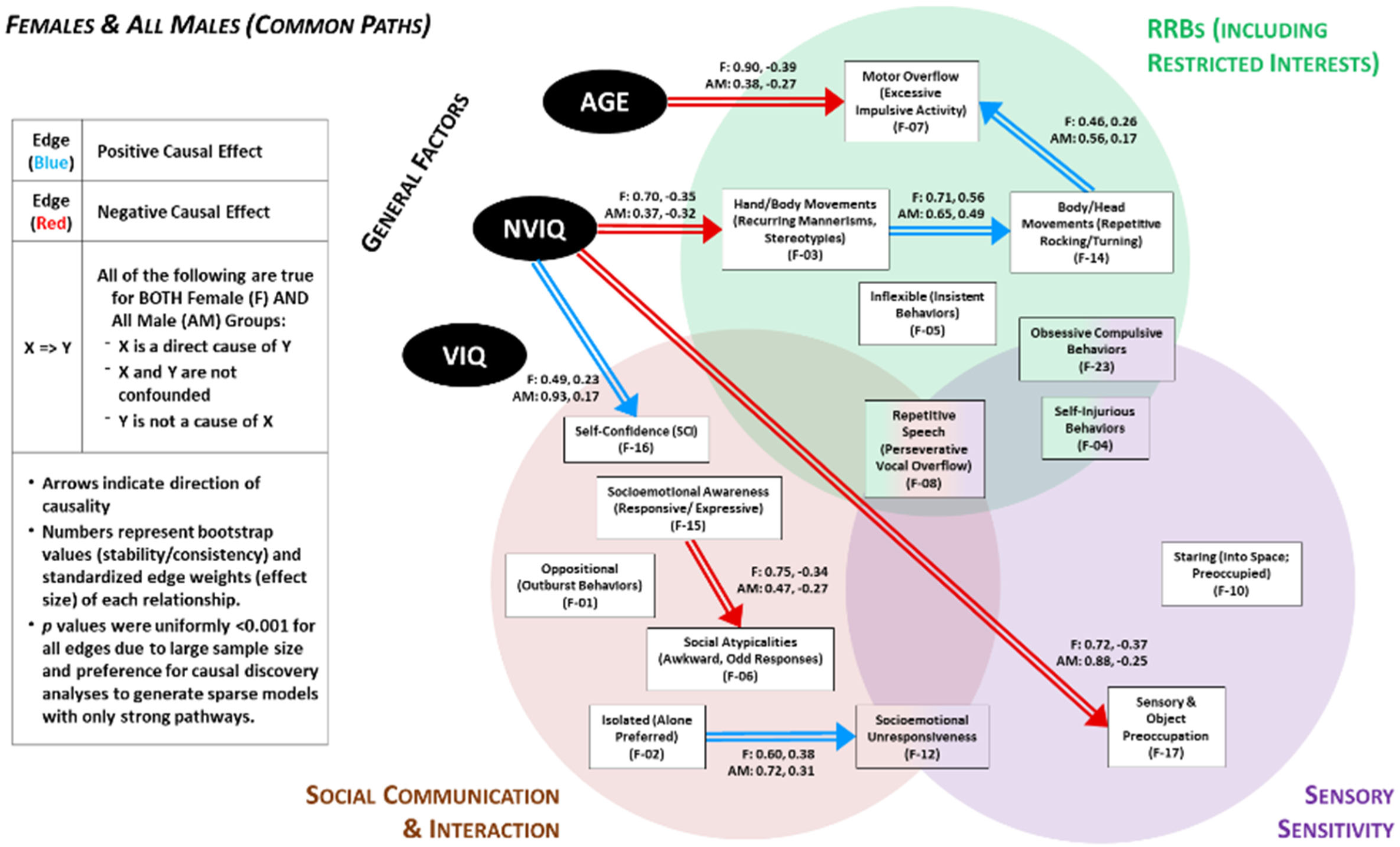
Directed Acyclic Graph suggested by the Greedy Fast Causal Inference (GFCI) causal discovery algorithm. Double arrows depict causal relations between factors that were common to both Female and All Male groups.

We also found associations between motor-related constructs such that more Hand/Body Movements (Recurring Mannerisms, Stereotypies) were linked to more Body/Head Movements (Repetitive Rocking/Turning) (F: 0.71, 0.56; MM: 0.63, 0.52). Increasing Motor Overflow (Excessive Impulsive Activity) was related to more Oppositional (Outburst Behaviors) acitivity (F: 0.50, 0.22; MM: 0.58, 0.31), which was downstream associated to an increase in Self-Injurious Behaviors (F: 0.56, 0.28; MM: 0.37, 0.25).

Variables related to social behavior both linked together and with compulsivity. Greater Socioemotional Awareness (Responsive/Expressive) was associated with a decrease in Social Atypicalities (Awkward, Odd Responses) (F: 0.75, -0.34; MM: 0.34, -0.26). An increase in Socioemotional Unresponsiveness related to an increase in Inflexible (Insistent Behaviors) (F: 0.37, 0.22; MM: 0.53, 0.31) and increasing Inflexible (Insistent Behaviors) related to an Increase in Obsessive Compulsive Behaviors (F: 0.66, 0.42; MM: 0.64, 0.52). Additionally, being more Isolated (Alone Preferred) related to more Socioemotional Unresponsiveness (F: 0.60, 0.38; MM: 0.72, 0.39) and increased Staring (into Space; Preoccupied) (F: 0.54, 0.32; MM: 0.62, 0.52).

**Figure 2.**
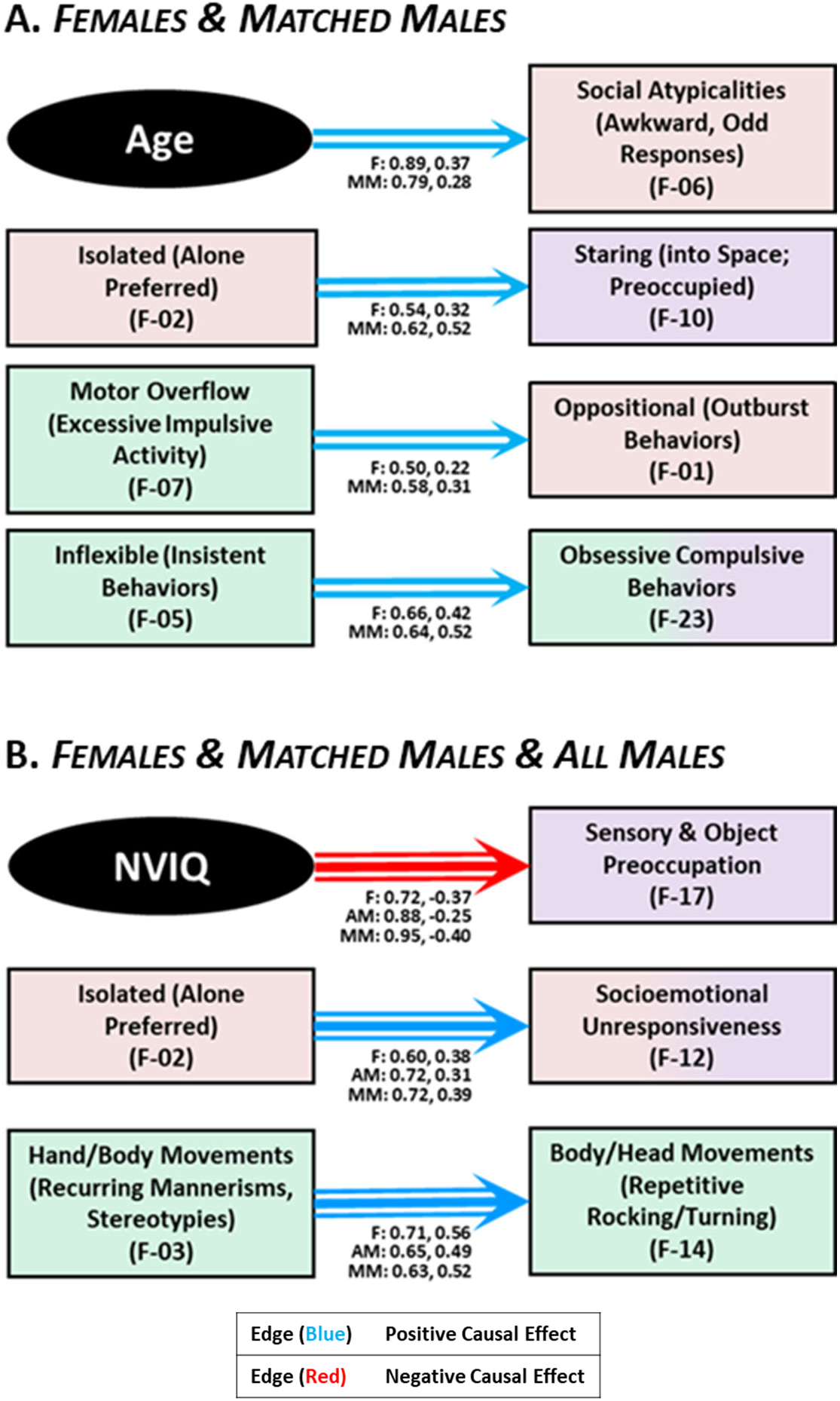
Causal connections between variables present in **(A.)** Female AND Matched Male groups and **(B.)** Females, Matched Males, AND All Male groups.

### FEMALES

For females in our dataset, CDA analysis yielded multiple directed relationships connecting general factors of AGE and IQ (NVIQ, VIQ) with clinical features (EFA-derived factors). Consistent with clinical observations of development, increasing AGE was related to decreasing Motor Overflow (Excessive Impulsive Activity) (F: 0.90, -0.39), increasing Social Atypicalities (Awkward, Odd Responses) (F: 0.89, 0.37), and increasing Socioemotional Unresponsiveness (F: 0.68, 0.22). Also consistent with established clinical observations, lower NVIQ was associated with an increase in Hand/Body Movements (Recurring Mannerisms, Stereotypies) (F: 0.72, -0.35) and more Sensory & Object Preoccupation (F: 0.72, -0.37). Lower VIQ was related to higher levels of Repetitive Speech (Perseverative Vocal Overflow) (F: 0.61, -0.28) and reduced Socioemotional Awareness (Responsive / Expressive) (F: 0.53, 0.28). Increased Socioemotional Awareness (Responsive/Expressive) was related to decreases in Social Atypicalities (Awkward, Odd Responses) (F: 0.75, -0.34). Although clinicians often observe these patterns with age and cognitive capacity, our findings highlight nuanced distinctions in the contributions of NVIQ versus VIQ to ASD phenomenology in females (Figure 3).

**Figure 3.**
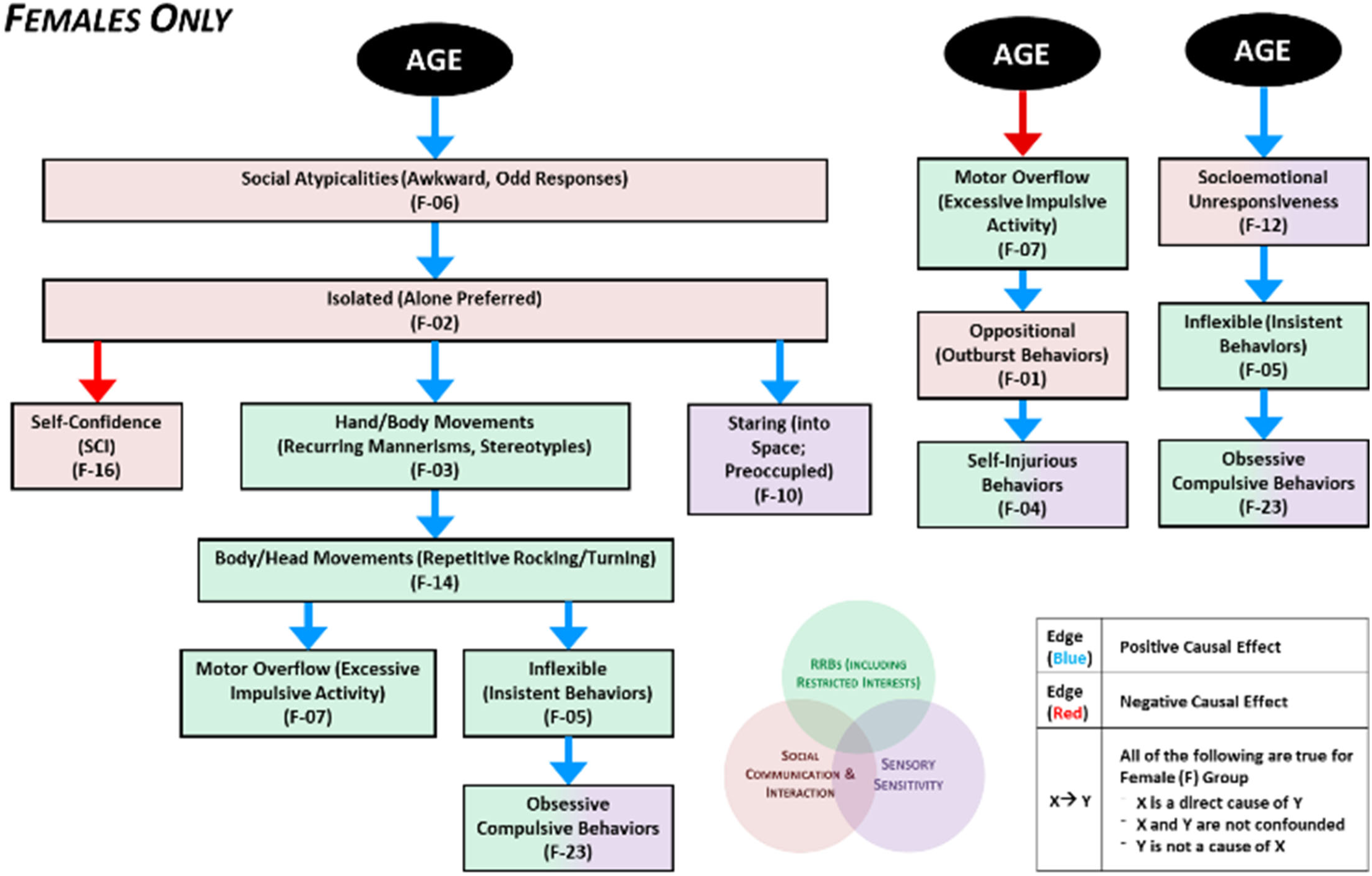
Causal pathways in females identified by CDA Analysis originating from AGE.

We also found multiple links between clinical features summarized in the EFA factors. First, we observed associations among overt repetitive, excessive movements, such that more Hand/Body Movements (Recurring Mannerisms, Stereotypies) were associated with more Body/Head Movements (Repetitive Rocking/Turning) (F: 0.71, 0.56)) and then increased Motor Overflow (Excessive Impulsive Activity), suggestive of convergent mechanistic relationships with neural systems underlying movement output and control. Interestingly, increased Motor Overflow (Excessive Impulsive Activity) was linked to an increase in Oppositional (Outburst Behaviors) activity (F: 0.50, 0.22) and, in turn, increased Self-Injurious Behaviors (F: 0.56, 0.28). This finding raises the possibility that motor systems play a central role in oppositional and self-injurious behavior in autistic females, rather than these behaviors being driven by an alternate process (e.g., emotion regulation). Second, we found relationships among compulsive behavioral tendencies (i.e., higher levels of Inflexible (Insistent Behaviors) were related to more Obsessive Compulsive Behaviors (F: 0.66, 0.42)). Finally, a preference for being Isolated (Alone Preferred) was associated with lower Self-Confidence (SCI) (F: 0.57, -0.31), more Socioemotional Unresponsiveness (F: 0.60, 0.38), and more Staring (into Space; Preoccupied) (F: 0.54, 0.32), suggesting that solitary preferences in females may drive subsequent social challenges.

### MATCHED MALES

In our subset of males matched to females for IQ and AGE, we observed some similar AGE associations wherein increasing AGE associated with decreasing in Motor Overflow (Excessive Impulsive Activity) (MM: 0.58, -0.22) and an increasing Social Atypicalities (Awkward, Odd Responses) (MM: 0.79, 0.28). In contrast to our female group, matched males did not show an association between increased AGE and increasing Socioemotional Unresponsiveness, suggesting putative sex-divergent trajectories in how emotion is expressed in social contexts, or in how autistic individuals are perceived to express such emotions (e.g., where socially-determined expectations for emotion expression differ by sex/gender).

Similar to females, the matched male group showed that lower NVIQ was related to increased Sensory & Object Preoccupation (MM: 0.95, -0.40). However, reduced Socioemotional Awareness (Responsive/Expressive) with lower NVIQ (MM: 0.64, 0.33) in matched males, whereas it was related to lower VIQ in females, pointing to the possibility that language and verbal abilities contribute differently to social functioning in autism based on sex, societal gender role expectations, or a combination of these factors.

Variables related to excess movement were associated with one another in matched males, as they were in females. Higher levels of Motor Overflow (Excessive Impulsive Activity) were associated with an Increase in Oppositional (Outburst Behaviors) (MM: 0.58, 0.31), and an increase in Hand/Body Movements (Recurring Mannerisms, Stereotypies) was linked to an increase in Body/Head Movements (Repetitive Rocking/Turning) (MM: 0.63, 0.52). Unlike females, data in MM did not show associations between motor-related processes and oppositional and self-injurious behaviors.

Similar to the female group, a greater preference for being Isolated (Alone Preferred) was accompanied by more Staring (into Space; Preoccupied) (MM: 0.62, 0.52) and increased Socioemotional Unresponsiveness (MM: 0.72, 0.39). An increase in Socioemotional Unresponsiveness was further related to an increase in Inflexible (Insistent Behaviors) (MM: 0.53, 0.31), and an increase in Inflexible (Insistent Behaviors) led to an increase in Obsessive Compulsive Behaviors (MM: 0.64, 0.52). Interestingly, males did not show the association between isolation and lower self-confidence that we observed in females, suggesting that gender or sex-related differences may shape how social experiences contribute to an autistic person’s sense of self.

## DISCUSSION

For our investigations, we first used exploratory factor analysis (EFA) to summarize clinical features across multiple validated assessments and then implemented causal discovery analyses (CDA) to model the structure and relationships between factors subserving general autism features (RRBs, SCI, Sens). CDA is a particularly powerful, data-driven tool that can detect directional influences between factors of interest while considering latent variables and their potentially confounding effects in the greater network. Our network theory-based approach discerned sex-biased, directional relationships between AGE, IQ, and clinical features, demonstrating the potential of using CDA to unearth phenotypically defined subtypes that will provide insight into the etiology of ASD and inform therapeutic targeting.

In order to examine how cognitive ability may impact symptom presentation, we considered the influence of verbal and non-verbal IQ in our sex/gender models of ASD. Prior research has suggested that cognitive profiles are affected by an individual’s pattern of performance in verbal and nonverbal reasoning - such that an IQ “discrepancy” or “split,” (e.g., NVIQ > VIQ or VIQ >NVIQ) may serve as a potential autism-related phenotype (Black, Wallace, Sokoloff, & Kenworthy, 2009; Chapman et al., 2010); sex- and age-based differences in cognitive discrepancy profiles have also been reported in ASD (Ankenman, Elgin, Sullivan, Vincent, & Bernier, 2014; Johnson et al., 2021). In an effort to control for IQ discrepancies that often occur when comparing autistic males and females, we included a subgroup of male participants (MM) matched individually to the smaller female sample by IQ and AGE. Matching procedures yielded a slightly younger male subgroup (MM: Age = 8.1 ± 3.2 years; FSIQ = 75.65 ± 27.13) than female group (F: Age = 9.2 ± 3.7 years; FSIQ = 76.29 ± 27.83); this may be attributed to the lower FSIQ (*p* < .01) overall in the female group than the larger male group (AM: Age = 8.9 ± 3.5 years; FSIQ = 82.01 ± 27.55). As such younger males were algorithmically selected to better match for FSIQ scores. Additionally, our sampling indicates that that males may show a larger NVIQ > VIQ split while female NVIQ and VIQ scores are more concordant.

Notably, while both and AGE and NVIQ were found to be causal ancestors of key factors in all three subgroups (F, AM, MM), VIQ was only an upstream factor for causal paths in the female group. In females, higher VIQ was related to less Repetitive Speech (Perseverative Vocal Overflow) and more Socioemotional Awareness (Responsive/Expressive). For all males and females, NVIQ was directly linked to factors in each domain, such that higher NVIQ associated with more Self-Confidence (SCI), reduced Hand/Body Movements (Recurring Mannerisms, Stereotypies) (RRBs), and less Sensory & Object Preoccupation (Sens). The presence of these common pathways suggest a more sex-independent role for NVIQ than VIQ in ASD. Further, while AGE showed a causal influence on both sexes, the variable originated more paths in females, suggesting more age-dependent related presentations and a more dynamic developmental pathway in female adolescence.

**Figure 4.**
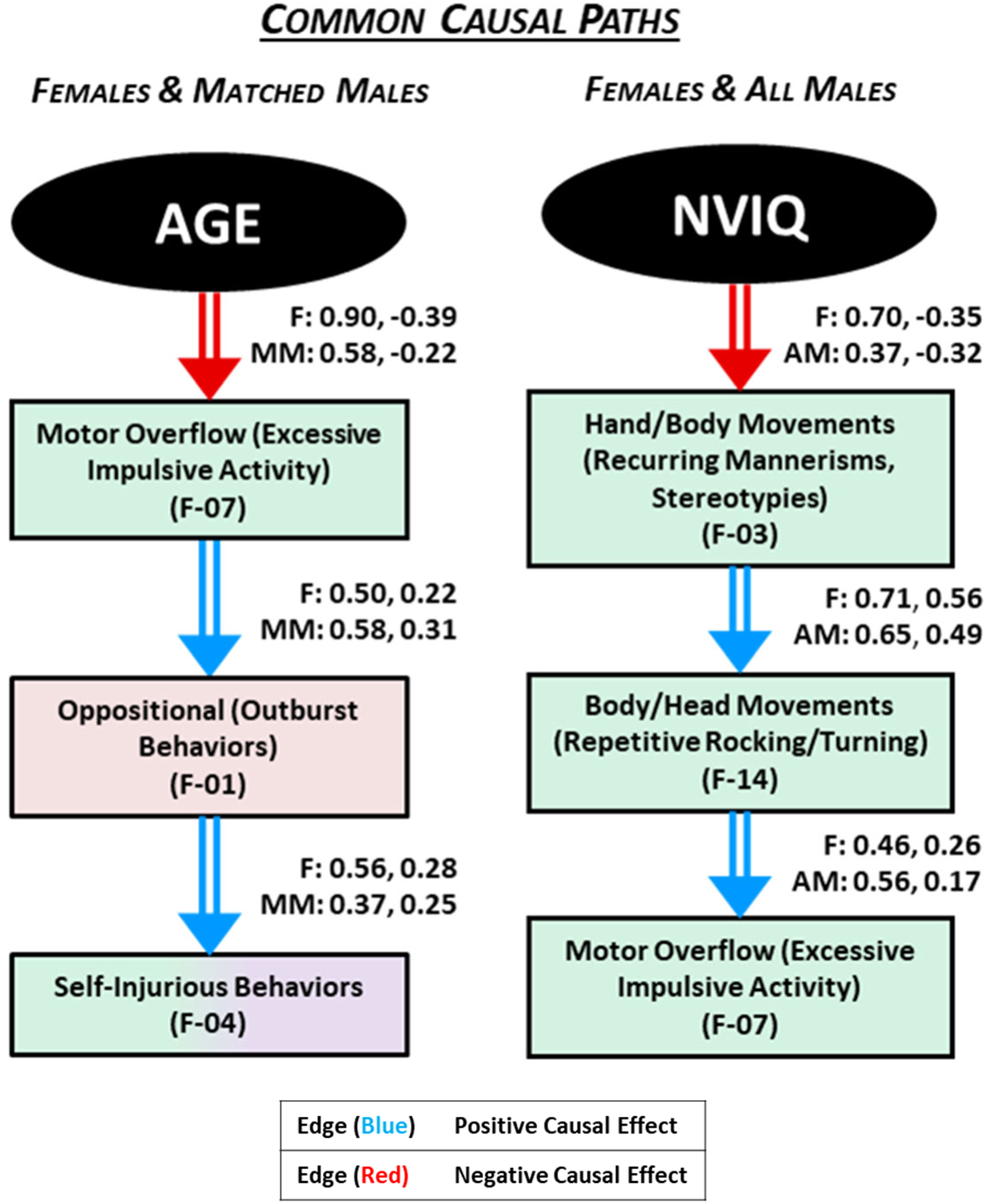
Common causal relationships originating from AGE for Females & Matched Males and from NVIQ for Females & All Males.

The CDA results and directional relationships align with our ‘real-world’ observations of how clinical presentations vary with IQ and change over time with autistic children. Differing and distinct relationships were found in subgroups: in our female sample (but not MM), increasing AGE was directly related to increased Socioemotional Unresponsiveness; in our matched male group (but not F) group we found that higher NVIQ was directly related to more Socioemotional Unresponsiveness. We consider that that these converging pathways highlight sex/gender differences in sociocultural expectations and phenotypic outcomes. For example, during the critical transitional periods of adolescence and adulthood, youth are exposed to more varied social situations that require complex interpersonal navigation. Females, who had been more successful at camouflaging with peers, may then struggle adjusting to the nuanced social requirements associated with increasing age. In contrast, societal expectations may penalize social disengagement less in males than in females, resulting in more stable observations of responsiveness with age. The association of higher NVIQ with increased Socioemotional Unresponsiveness in the matched male group may reflect the greater NVIQ > VIQ discrepancy in our male sample; verbal ability is inexorably linked to social communication and would be impacted by the lower VIQ accompanied by higher NVIQ.

**Figure 5.**
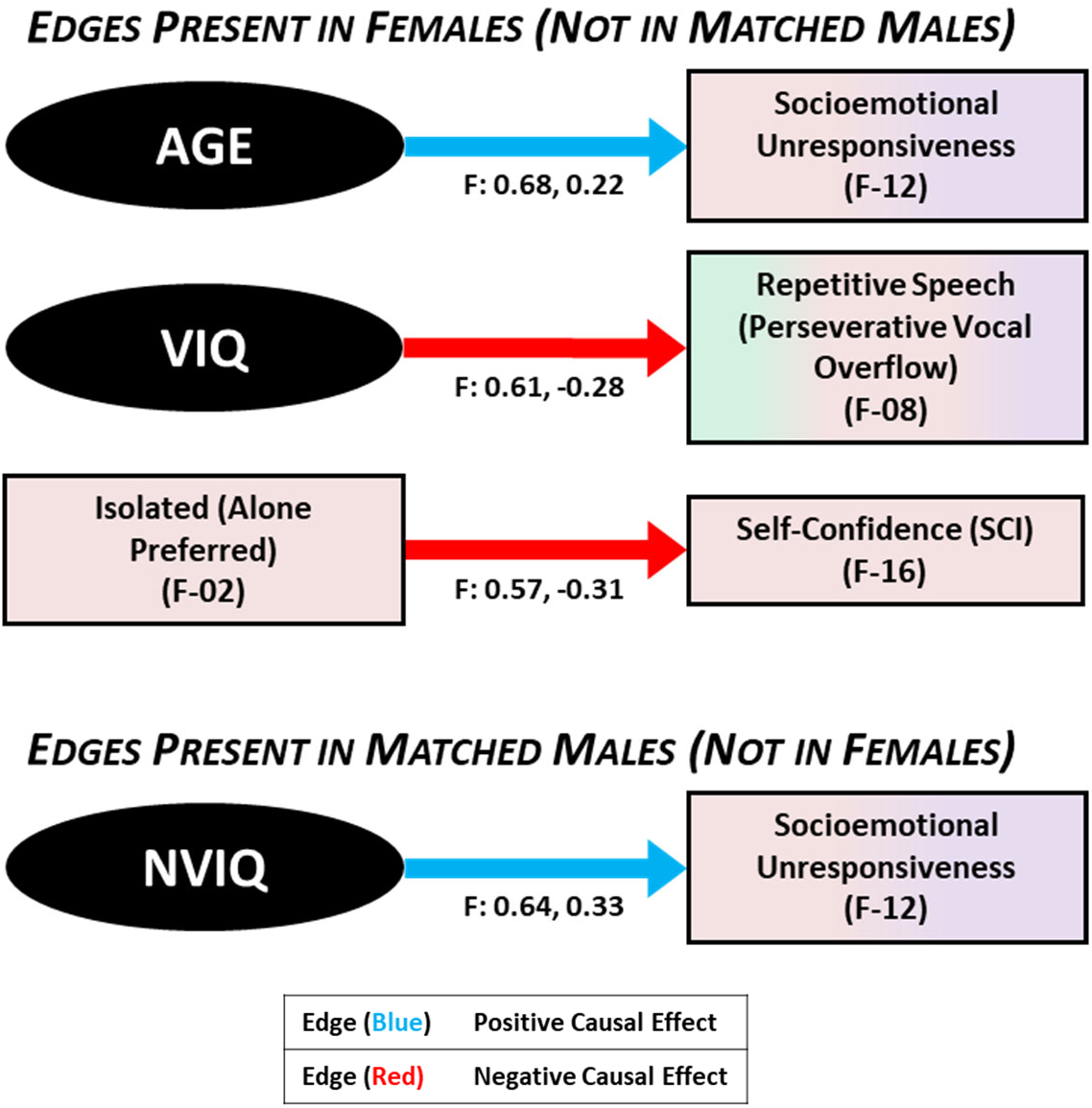
Causal connections present in Females (but not Matched Males) and in Matched Males (but not Females).

Investigating causal pathways between the EFA-derived factors yielded patterns that may enable us to better conceptualize heterogeneity in autism symptom expression across sexes. Further, the mapping of causal influences on ASD-related outcomes may identify targets for intervention at nodes with mediating or indirect causal effects in the pathways. For example, in both our female and matched male groups, we found that an an increase in Self-Injurious Behaviors was indirectly caused by an increase in Motor Overflow (Excessive Impulsive Activity) mediated through an increase in Oppositional (Outburst Behaviors). Our analysis indicates that mitigating behaviors in the RRB domain may lead to a reduction in both oppositional and self-injurious behaviors.

### Limitations

To our knowledge, our analyses represent the first implementation of this causal discovery approach in a large sample of well-characterized youth with ASD. However, we must address limitations in our methodology and dataset. First, given the dynamic nature of the autism field of study, wherein diagnostic criteria, standards, and biases are subject to scrutiny and modification, the SSC v15.3 dataset reflects the state of the field during the period of collection. For example, new to the DSM-5 definition of autism was the inclusion of “hyper or hypo reactivity to sensory input or unusual interest in sensory aspects of the environment” as one of the four restricted/repetitive behavior features defined as atypical sensory processing (American Psychiatric Association, 2013). Consequently, earlier assessments may have been lacking in sensory domain items. Relatedly, the available dataset also inexorably reflects the ongoing sex/gender-biased diagnostic discrepancies discussed earlier. Hence, the male to female ratio of our included sample is closer to 6 males:1 female, likely reflecting the under-diagnosis of autistic girls who were therefore not recruited to contribute to the Simons Simplex Collection studies. In response to these concerns, and with the goal of better understanding differences and similarities in male vs. female representations of ASD without the confounding, we created a sample of males matched individually to all available females to offset confounds that may be introduced by age and FSIQ variability. Finally, we acknowledge the inherent limitations of parent/caregiver/other-report measures as a subjective source of data. However, given the development age group of interest (Age < 18 years), questionnaire data from validated measures are the most feasible approach to better understand these vulnerable populations.

### Conclusions

To our knowledge, this is the first study to examine sex differences while modeling direct causal pathways between AGE, IQ, and ASD characteristics across RRB, SCI, and Sens domains in a large sample of autistic youth. By implementing an analytical methodology originally designed for causal discovery from observational data broadly, we unveiled sex-specific directional relationships between multiple clinically-relevant variables. Our findings highlight potential applications for CDA as a means to understand mechanisms of ASD symptomatology, including latent influences on phenotypic outcomes. Further research may impact downstream outcomes (e.g., self-injurious or oppositional behaviors) by illuminating upstream targets for intervention in discovered causal pathways.

## Data Availability

Approved researchers can obtain the Simon Simplex Collection (SSC) dataset described in this study (v15) by applying at https://base.sfari.org.

## AUTHORS’ CONTRIBUTIONS

- SF – Data Curation, Formal Analysis, Methodology, Writing - Original Draft, Writing - Review & Editing, Project Administration, Visualization
- AT – Conceptualization, Methodology, Writing - Original Draft, Writing - Review & Editing, Visualization
- ER – Formal Analysis, Methodology, Writing - Review & Editing
- CC – Clinical Interpretation, Writing - Review & Editing
- NG – Conceptualization, Writing - Review & Editing
- EK – Formal Analysis, Methodology, Writing - Review & Editing
- SM – Conceptualization, Methodology, Writing - Review & Editing, Supervision
- SJ – Conceptualization, Methodology, Project Funding, Clinical Interpretation, Writing - Review & Editing, Supervision

## ACKNOWLEDGEMENTS

We thank the NeuroPlasticity Research in Support of Mental Health (NeuroPRSMH) group at the University of Minnesota for helpful discussions. We are grateful to Megan Dubois and Noor Christava for their assistance with data organization. We also thank the families who contributed to the Simons Simplex Collection. Approved researchers can obtain the SSC dataset described in this study (v15) by applying at https://base.sfari.org.

## Footnotes

1 We use the term sex/gender to acknowledge the overlap between biological characteristics (sex) and socially constructed attributes and expectations (gender) as proposed in (Springer et al. 2012).

