## Supplemental Figure 1 for "Integrating Causal Discovery and Clinically-Relevant Insights to Explore Directional Relationships between Autistic Features, Sex at Birth, and Cognitive Abilities"

### **SUPPLEMENTAL MATERIALS**

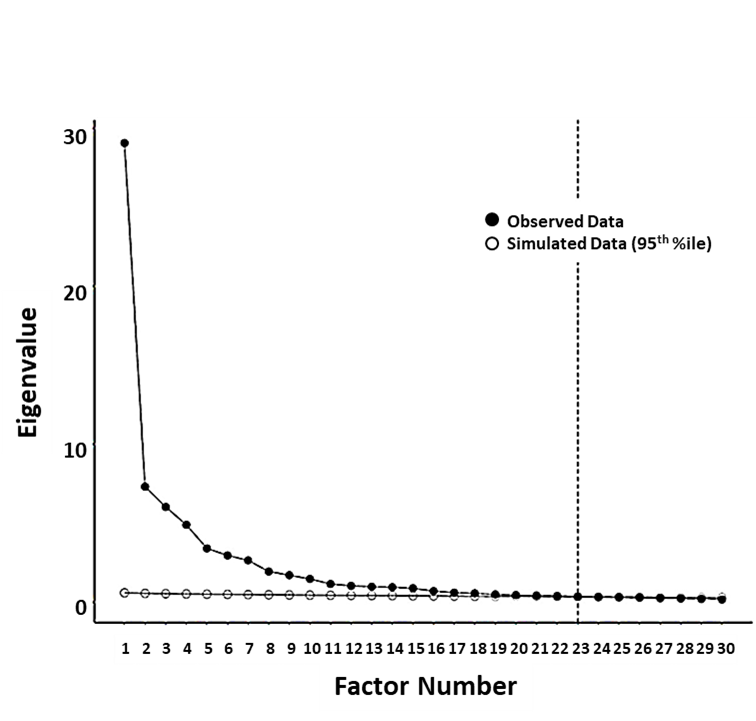

**Figure S1.** Depicts eigenvalues and 23 factors meeting EFA criteria.

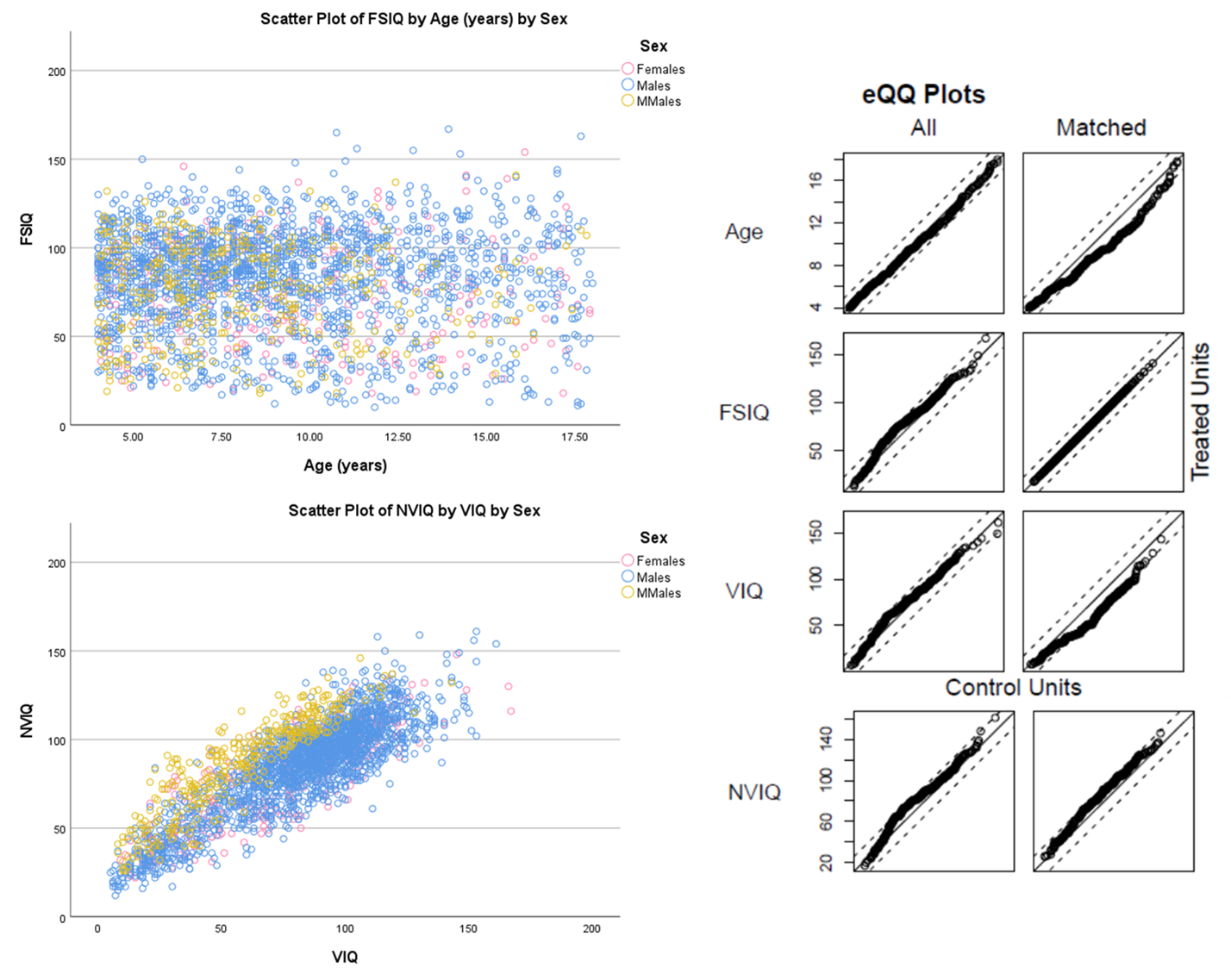

**Figure S2.** Matched Male Subgroup was generated using exact matching for FSIQ and nearest matching for AGE, VIQ, and NVIQ variables.
