## Supplemental Table 1 for "Integrating Causal Discovery and Clinically-Relevant Insights to Explore Directional Relationships between Autistic Features, Sex at Birth, and Cognitive Abilities"

### SUPPLEMENTAL MATERIALS

**Table S1.** All Exploratory Factor Analysis (EFA) derived factors. Bolding indicates factors that were excluded from final analyses after consensus review by clinical experts.

| Factor # | Label Name | Assessments |
|---|---|---|
| F-01 | Oppositional (Outburst Behaviors) | ABC |
| F-02 | Isolated (Alone Preferred) | ABC; SRS |
| F-03 | Hand/Body Movements (Recurring Mannerisms, Stereotypies) | ABC; RBS-R; SRS |
| F-04 | Self-Injurious Behaviors | ABC; RBS-R |
| F-05 | Inflexible (Insistent Behaviors) | RBS-R; SRS |
| F-06 | Social Atypicalities (Awkward, Odd Responses) | SRS |
| F-07 | Motor Overflow (Excessive Impulsive Activity) | ABC; RBS-R |
| F-08 | Repetitive Speech (Perseverative Vocal Overflow) | ABC; RBS-R; SRS |
| **F-09** | **Decreased Conversational Comprehension** | **SRS** |
| F-10 | Staring (into Space; Preoccupied) | ABC; SRS |
| **F-11** | **Tantrums** | **ABC** |
| F-12 | Socioemotional Unresponsiveness | ABC; SRS |
| **F-13** | **Inflexibility Associated with Social Stress** | **SRS** |
| F-14 | Body/Head Movements (Repetitive Rocking/Turning) | ABC; RBS-R |
| F-15 | Socioemotional Awareness (Responsive/Expressive) | SRS |
| F-16 | Self-Confidence (Social Communication & Interaction) | SRS |
| F-17 | Sensory & Object Preoccupation | RBS-R; SRS |
| **F-18** | **Restlessness** | **ABC** |
| **F-19** | **Temperamental/Irritable** | **ABC** |
| **F-20** | **Restricted Interests/Discussion Topics** | **RBS-R; SRS** |
| **F-21** | **Self-Injurious Behaviors (only characterized by RBS-R)** | **RBS-R** |
| **F-22** | **Teased** | **SRS** |
| F-23 | Obsessive Compulsive Behaviors | RBS-R |

**Assessments:** Aberrant Behavior Checklist-Community Version (ABC-CV); Repetitive Behavior Scale-Revised (RBS-R); Social Responsiveness Scale (SRS).
**Domains:** Social Communication & Interaction (SCI); Restricted, Repetitive Patterns of Behavior, Interests, or Activities (RRBs); Sensory Sensitivity (Sens)
