## Supplemental Table 2 for "Integrating Causal Discovery and Clinically-Relevant Insights to Explore Directional Relationships between Autistic Features, Sex at Birth, and Cognitive Abilities"

**Table S2.** Causal Discovery Analysis Bootstrap Re-sampling Results for (A.) Females, (B.) All Males, and (C.) Matched Males.

### A. FEMALES

| | Node 1 | Interaction | Node 2 | Ensemble | No Edge | --> | <-- | --> | <-- | --> | <-- | o-> | <-o | o-o | --- |
|---|---|---|---|---|---|---|---|---|---|---|---|---|---|---|---|
| 1 | AGE | o-> | F-07: Motor Overflow (Excessive Impulsive Activity) | 0.90 | 0.00 | 0.00 | | | | 0.00 | | 0.90 | | | 0.09 |
| 2 | AGE | o-> | F-06: Social Atypicalities (Awkward, Odd Responses) | 0.89 | 0.01 | 0.01 | | 0.00 | | 0.00 | | 0.89 | | | 0.09 |
| 3 | F-15: Socioemotional Awareness (Responsive/Expressive) | --> | F-06: Social Atypicalities (Awkward, Odd Responses) | 0.75 | 0.03 | | | 0.02 | 0.01 | 0.75 | 0.10 | 0.07 | | | 0.02 |
| 4 | F-17: Sensory & Object Preoccupation | <-o | NVIQ | 0.72 | 0.23 | | 0.03 | | 0.00 | | 0.02 | | 0.72 | | |
| 5 | F-14: Body/Head Movements (Repetitive Rocking/Turning) | <-- | F-03: Hand/Body Movements (Recurring Mannerisms, Stereotypies) | 0.71 | 0.00 | | | 0.01 | 0.06 | 0.21 | 0.71 | 0.00 | 0.00 | 0.01 | |
| 6 | F-03: Hand/Body Movements (Recurring Mannerisms, Stereotypies) | <-o | NVIQ | 0.70 | 0.27 | | 0.02 | | 0.01 | | 0.00 | | 0.70 | | 0.00 |
| 7 | AGE | o-> | F-12: Socioemotional Unresponsiveness | 0.68 | 0.25 | | | 0.01 | | 0.06 | | 0.68 | | | 0.00 |
| 8 | F-23: Obsessive Compulsive Behaviors | <-- | F-05: Inflexible (Insistent Behaviors) | 0.66 | 0.00 | | | 0.03 | 0.03 | 0.26 | 0.66 | 0.00 | 0.01 | 0.01 | 0.00 |
| 9 | F-08: Repetitive Speech (Perseverative Vocal Overflow) | <-o | VIQ | 0.61 | 0.35 | | 0.01 | | 0.00 | | 0.01 | | 0.61 | | 0.02 |
| 10 | F-12: Socioemotional Unresponsiveness | <-- | F-02: Isolated (Alone Preferred) | 0.60 | 0.00 | | 0.00 | 0.03 | 0.03 | 0.32 | 0.60 | | 0.01 | 0.01 | 0.00 |
| 11 | F-16: Self-Confidence (SCI) | <-- | F-02: Isolated (Alone Preferred) | 0.57 | 0.20 | | | 0.01 | 0.07 | 0.09 | 0.57 | 0.01 | 0.02 | 0.00 | 0.03 |
| 12 | F-01: Oppositional (Outburst Behaviors) | --> | F-04: Self-Injurious Behaviors | 0.56 | 0.04 | | | 0.01 | 0.02 | 0.56 | 0.37 | 0.00 | 0.00 | 0.00 | |
| 13 | F-10: Staring (into Space; Preoccupied) | <-- | F-02: Isolated (Alone Preferred) | 0.54 | 0.09 | | | 0.01 | 0.12 | 0.15 | 0.54 | 0.04 | 0.02 | 0.03 | 0.01 |
| 14 | F-15: Socioemotional Awareness (Responsive/Expressive) | <-o | VIQ | 0.53 | 0.45 | | 0.01 | | 0.00 | | 0.01 | | 0.53 | | 0.00 |
| 15 | F-01: Oppositional (Outburst Behaviors) | <-- | F-07: Motor Overflow (Excessive Impulsive Activity) | 0.50 | 0.39 | | | 0.00 | 0.06 | 0.04 | 0.50 | | | | |
| 16 | F-16: Self-Confidence (SCI) | <-o | NVIQ | 0.49 | 0.49 | | 0.00 | | | | | | 0.49 | | 0.02 |
| 17 | F-14: Body/Head Movements (Repetitive Rocking/Turning) | --> | F-07: Motor Overflow (Excessive Impulsive Activity) | 0.46 | 0.49 | | | 0.02 | 0.00 | 0.46 | 0.03 | 0.01 | | | |
| 18 | F-02: Isolated (Alone Preferred) | <-- | F-06: Social Atypicalities (Awkward, Odd Responses) | 0.43 | 0.41 | | | 0.02 | 0.03 | 0.11 | 0.43 | 0.00 | | | 0.00 |
| 19 | F-14: Body/Head Movements (Repetitive Rocking/Turning) | --> | F-05: Inflexible (Insistent Behaviors) | 0.40 | 0.38 | | | 0.11 | 0.03 | 0.40 | 0.08 | 0.00 | 0.00 | 0.00 | 0.00 |
| 20 | F-01: Oppositional (Outburst Behaviors) | <-- | F-05: Inflexible (Insistent Behaviors) | 0.37 | 0.44 | 0.00 | | 0.01 | 0.04 | 0.14 | 0.37 | | 0.00 | | |
| 21 | F-12: Socioemotional Unresponsiveness | --> | F-05: Inflexible (Insistent Behaviors) | 0.37 | 0.46 | | | 0.08 | 0.01 | 0.37 | 0.07 | 0.00 | 0.00 | 0.00 | 0.00 |
| 22 | F-07: Motor Overflow (Excessive Impulsive Activity) | <-o | VIQ | 0.36 | 0.64 | | | | 0.00 | | 0.01 | | 0.36 | | 0.00 |
| 23 | F-04: Self-Injurious Behaviors | <-o | VIQ | 0.35 | 0.64 | | 0.00 | | 0.00 | | 0.01 | | 0.35 | | 0.00 |
| 24 | F-02: Isolated (Alone Preferred) | --> | F-03: Hand/Body Movements (Recurring Mannerisms, Stereotypies) | 0.32 | 0.41 | | | 0.04 | 0.04 | 0.32 | 0.16 | 0.01 | 0.00 | 0.00 | 0.01 |
| 25 | F-15: Socioemotional Awareness (Responsive/Expressive) | <-- | F-17: Sensory & Object Preoccupation | 0.30 | 0.53 | | | 0.03 | 0.05 | 0.08 | 0.30 | 0.01 | 0.00 | 0.01 | |
| 26 | F-23: Obsessive Compulsive Behaviors | <-- | F-07: Motor Overflow (Excessive Impulsive Activity) | 0.25 | 0.62 | | | 0.00 | 0.07 | 0.06 | 0.25 | | | | |
| 27 | F-17: Sensory & Object Preoccupation | --> | F-23: Obsessive Compulsive Behaviors | 0.24 | 0.50 | | | 0.05 | 0.00 | 0.24 | 0.19 | | 0.01 | | 0.00 |
| 28 | F-10: Staring (into Space; Preoccupied) | <-- | F-03: Hand/Body Movements (Recurring Mannerisms, Stereotypies) | 0.22 | 0.57 | | | 0.01 | 0.06 | 0.12 | 0.22 | 0.02 | 0.01 | | |
| 29 | F-14: Body/Head Movements (Repetitive Rocking/Turning) | <-- | F-17: Sensory & Object Preoccupation | 0.20 | 0.60 | | | 0.02 | 0.05 | 0.13 | 0.20 | | 0.00 | 0.00 | |
| 30 | F-14: Body/Head Movements (Repetitive Rocking/Turning) | --> | F-04: Self-Injurious Behaviors | 0.17 | 0.69 | | | 0.08 | 0.01 | 0.17 | 0.05 | 0.00 | 0.00 | 0.00 | 0.00 |
| 31 | F-14: Body/Head Movements (Repetitive Rocking/Turning) | --> | F-08: Repetitive Speech (Perseverative Vocal Overflow) | 0.17 | 0.76 | | | 0.01 | 0.01 | 0.17 | 0.05 | 0.00 | | | |
| 32 | F-17: Sensory & Object Preoccupation | <-o | VIQ | 0.17 | 0.83 | | | | 0.00 | | 0.00 | | 0.17 | | |
| 33 | AGE | o-> | F-16: Self-Confidence (SCI) | 0.17 | 0.82 | | | 0.01 | | 0.01 | | 0.17 | | | 0.00 |
| 34 | AGE | o-> | F-04: Self-Injurious Behaviors | 0.16 | 0.82 | | | 0.01 | | 0.01 | | 0.16 | | | 0.01 |
| 35 | F-16: Self-Confidence (SCI) | <-- | F-08: Repetitive Speech (Perseverative Vocal Overflow) | 0.16 | 0.67 | | | 0.00 | 0.01 | 0.14 | 0.16 | 0.01 | 0.00 | | 0.01 |
| 36 | F-07: Motor Overflow (Excessive Impulsive Activity) | --> | F-08: Repetitive Speech (Perseverative Vocal Overflow) | 0.16 | 0.78 | | | 0.01 | | 0.16 | 0.05 | | | | |
| 37 | F-10: Staring (into Space; Preoccupied) | <-- | F-06: Social Atypicalities (Awkward, Odd Responses) | 0.15 | 0.67 | | | 0.01 | 0.05 | 0.10 | 0.15 | 0.01 | | 0.00 | |
| 38 | F-15: Socioemotional Awareness (Responsive/Expressive) | --> | F-16: Self-Confidence (SCI) | 0.15 | 0.67 | | | 0.07 | 0.00 | 0.15 | 0.10 | 0.00 | 0.00 | 0.00 | |
| 39 | F-12: Socioemotional Unresponsiveness | --> | F-01: Oppositional (Outburst Behaviors) | 0.15 | 0.78 | | | 0.03 | 0.00 | 0.15 | 0.04 | 0.00 | 0.00 | | 0.00 |
| 40 | F-08: Repetitive Speech (Perseverative Vocal Overflow) | <-o | NVIQ | 0.14 | 0.84 | | 0.00 | | | | 0.00 | | 0.14 | | 0.02 |
| 41 | F-10: Staring (into Space; Preoccupied) | --> | F-08: Repetitive Speech (Perseverative Vocal Overflow) | 0.13 | 0.83 | | | 0.01 | 0.00 | 0.13 | 0.03 | 0.01 | | | |
| 42 | F-23: Obsessive Compulsive Behaviors | --> | F-08: Repetitive Speech (Perseverative Vocal Overflow) | 0.13 | 0.76 | | | 0.01 | 0.01 | 0.13 | 0.09 | 0.00 | | | |
| 43 | F-17: Sensory & Object Preoccupation | --> | F-03: Hand/Body Movements (Recurring Mannerisms, Stereotypies) | 0.12 | 0.71 | | | 0.03 | 0.02 | 0.12 | 0.08 | 0.01 | 0.01 | 0.01 | |
| 44 | F-12: Socioemotional Unresponsiveness | <-o | VIQ | 0.10 | 0.89 | | | | | | | | 0.10 | | 0.01 |
| 45 | F-04: Self-Injurious Behaviors | <-o | NVIQ | 0.09 | 0.91 | | | | | | | | 0.09 | | |
| 46 | F-17: Sensory & Object Preoccupation | <-- | F-05: Inflexible (Insistent Behaviors) | 0.09 | 0.83 | | | 0.03 | 0.00 | 0.05 | 0.09 | | 0.00 | | |
| 47 | F-16: Self-Confidence (SCI) | <-o | VIQ | 0.09 | 0.89 | | 0.01 | | 0.00 | | 0.00 | | 0.09 | | 0.02 |
| 48 | F-10: Staring (into Space; Preoccupied) | <-- | F-14: Body/Head Movements (Repetitive Rocking/Turning) | 0.08 | 0.87 | | | 0.00 | 0.02 | 0.02 | 0.08 | | 0.00 | 0.00 | 0.00 |
| 49 | F-03: Hand/Body Movements (Recurring Mannerisms, Stereotypies) | <-o | VIQ | 0.08 | 0.92 | | | | | | 0.00 | | 0.08 | | |
| 50 | F-12: Socioemotional Unresponsiveness | --> | F-16: Self-Confidence (SCI) | 0.06 | 0.92 | | | 0.01 | | 0.06 | 0.00 | | | | |
| 51 | F-01: Oppositional (Outburst Behaviors) | <-- | F-06: Social Atypicalities (Awkward, Odd Responses) | 0.05 | 0.92 | | | 0.00 | 0.02 | 0.01 | 0.05 | | | | |
| 52 | F-10: Staring (into Space; Preoccupied) | --> | F-17: Sensory & Object Preoccupation | 0.05 | 0.89 | | | 0.01 | 0.02 | 0.05 | 0.02 | 0.01 | | | 0.00 |
| 53 | F-03: Hand/Body Movements (Recurring Mannerisms, Stereotypies) | --> | F-07: Motor Overflow (Excessive Impulsive Activity) | 0.05 | 0.90 | | | 0.01 | 0.00 | 0.05 | 0.04 | | | | |
| 54 | F-02: Isolated (Alone Preferred) | --> | F-05: Inflexible (Insistent Behaviors) | 0.04 | 0.89 | | | 0.03 | 0.01 | 0.04 | 0.02 | 0.00 | 0.00 | 0.00 | |
| 55 | F-04: Self-Injurious Behaviors | <-- | F-08: Repetitive Speech (Perseverative Vocal Overflow) | 0.04 | 0.92 | | | 0.00 | 0.00 | 0.03 | 0.04 | | | | |
| 56 | F-15: Socioemotional Awareness (Responsive/Expressive) | <-- | F-07: Motor Overflow (Excessive Impulsive Activity) | 0.04 | 0.94 | | | 0.00 | 0.01 | 0.00 | 0.04 | | | | |
| 57 | F-01: Oppositional (Outburst Behaviors) | <-- | F-08: Repetitive Speech (Perseverative Vocal Overflow) | 0.04 | 0.92 | | | 0.00 | 0.00 | 0.04 | 0.04 | | | | |
| 58 | F-03: Hand/Body Movements (Recurring Mannerisms, Stereotypies) | --> | F-08: Repetitive Speech (Perseverative Vocal Overflow) | 0.04 | 0.93 | | | 0.02 | | 0.04 | 0.01 | 0.00 | | | |
| 59 | F-03: Hand/Body Movements (Recurring Mannerisms, Stereotypies) | --> | F-05: Inflexible (Insistent Behaviors) | 0.04 | 0.92 | | | 0.02 | 0.00 | 0.04 | 0.02 | | 0.00 | 0.00 | |
| 60 | F-23: Obsessive Compulsive Behaviors | <-o | VIQ | 0.04 | 0.96 | | | | | | 0.00 | | 0.04 | | |
| 61 | AGE | o-> | F-02: Isolated (Alone Preferred) | 0.04 | 0.95 | 0.00 | | 0.00 | | 0.01 | | 0.04 | | | |
| 62 | F-17: Sensory & Object Preoccupation | --> | F-07: Motor Overflow (Excessive Impulsive Activity) | 0.04 | 0.93 | | | 0.00 | | 0.04 | 0.03 | | | | |
| 63 | F-15: Socioemotional Awareness (Responsive/Expressive) | <-- | F-02: Isolated (Alone Preferred) | 0.03 | 0.94 | | | 0.01 | 0.01 | 0.01 | 0.03 | | | | |
| 64 | AGE | o-> | F-05: Inflexible (Insistent Behaviors) | 0.03 | 0.97 | | | 0.00 | | 0.00 | | 0.03 | | | |
| 65 | F-06: Social Atypicalities (Awkward, Odd Responses) | <-- | F-08: Repetitive Speech (Perseverative Vocal Overflow) | 0.03 | 0.96 | | | 0.00 | 0.01 | 0.00 | 0.03 | | | | |
| 66 | F-02: Isolated (Alone Preferred) | --> | F-04: Self-Injurious Behaviors | 0.03 | 0.96 | | | 0.00 | | 0.03 | 0.01 | | | | |
| 67 | F-15: Socioemotional Awareness (Responsive/Expressive) | <-o | NVIQ | 0.03 | 0.97 | | 0.00 | | | | | | 0.03 | | |

| | | | | | | | | | | | | | | | |
|---|---|---|---|---|---|---|---|---|---|---|---|---|---|---|---|
| 68 | F-02: Isolated (Alone Preferred) | <-- | F-07: Motor Overflow (Excessive Impulsive Activity) | 0.03 | 0.97 | | | | 0.00 | | 0.03 | | | | |
| 69 | F-17: Sensory & Object Preoccupation | --> | F-04: Self-Injurious Behaviors | 0.03 | 0.95 | | | 0.03 | | 0.02 | 0.00 | | | | |
| 70 | F-23: Obsessive Compulsive Behaviors | --> | F-02: Isolated (Alone Preferred) | 0.03 | 0.96 | | | 0.01 | 0.01 | 0.03 | 0.01 | 0.00 | | 0.00 | |
| 71 | F-14: Body/Head Movements (Repetitive Rocking/Turning) | <-o | VIQ | 0.03 | 0.97 | | 0.00 | | | | | | 0.03 | | |
| 72 | F-15: Socioemotional Awareness (Responsive/Expressive) | --> | F-03: Hand/Body Movements (Recurring Mannerisms, Stereotypies) | 0.03 | 0.94 | | | 0.01 | 0.01 | 0.03 | 0.02 | | | | |
| 73 | F-15: Socioemotional Awareness (Responsive/Expressive) | <-- | F-01: Oppositional (Outburst Behaviors) | 0.03 | 0.95 | | | 0.01 | | 0.02 | 0.03 | | | | |
| 74 | F-10: Staring (into Space; Preoccupied) | --> | F-01: Oppositional (Outburst Behaviors) | 0.03 | 0.96 | | | 0.00 | 0.00 | 0.03 | 0.01 | 0.00 | | | |
| 75 | F-17: Sensory & Object Preoccupation | <-- | F-02: Isolated (Alone Preferred) | 0.02 | 0.97 | | | 0.00 | 0.00 | | 0.02 | | 0.00 | | 0.00 |
| 76 | F-16: Self-Confidence (SCI) | <-- | F-07: Motor Overflow (Excessive Impulsive Activity) | 0.02 | 0.98 | | | | | | 0.02 | | | | |
| 77 | F-12: Socioemotional Unresponsiveness | --> | F-23: Obsessive Compulsive Behaviors | 0.02 | 0.96 | | | 0.01 | | 0.02 | 0.01 | | | | |
| 78 | F-12: Socioemotional Unresponsiveness | <-- | F-14: Body/Head Movements (Repetitive Rocking/Turning) | 0.02 | 0.95 | | | 0.01 | 0.01 | 0.01 | 0.02 | 0.00 | | | |
| 79 | AGE | o-> | F-17: Sensory & Object Preoccupation | 0.02 | 0.98 | | | 0.00 | | 0.00 | | 0.02 | | | |
| 80 | F-16: Self-Confidence (SCI) | <-- | F-06: Social Atypicalities (Awkward, Odd Responses) | 0.02 | 0.97 | | | 0.00 | 0.00 | 0.00 | 0.02 | | | | |
| 81 | F-04: Self-Injurious Behaviors | --> | F-07: Motor Overflow (Excessive Impulsive Activity) | 0.02 | 0.98 | | | 0.00 | | 0.02 | 0.00 | 0.00 | | | |
| 82 | F-07: Motor Overflow (Excessive Impulsive Activity) | <-o | NVIQ | 0.02 | 0.98 | | | | | | | | 0.02 | | |
| 83 | F-05: Inflexible (Insistent Behaviors) | --> | F-08: Repetitive Speech (Perseverative Vocal Overflow) | 0.02 | 0.98 | | | 0.00 | | 0.02 | 0.00 | | | | |
| 84 | AGE | o-> | F-15: Socioemotional Awareness (Responsive/Expressive) | 0.01 | 0.98 | 0.00 | | | | 0.00 | | 0.01 | | | 0.00 |
| 85 | F-02: Isolated (Alone Preferred) | --> | F-08: Repetitive Speech (Perseverative Vocal Overflow) | 0.01 | 0.98 | | | 0.00 | | 0.01 | 0.00 | | | | 0.00 |
| 86 | AGE | o-> | F-14: Body/Head Movements (Repetitive Rocking/Turning) | 0.01 | 0.99 | | | | | | | 0.01 | | | 0.00 |
| 87 | F-16: Self-Confidence (SCI) | --> | F-04: Self-Injurious Behaviors | 0.01 | 0.99 | | | 0.00 | | 0.01 | | | | | |
| 88 | F-10: Staring (into Space; Preoccupied) | <-o | VIQ | 0.01 | 0.99 | | | | | | 0.00 | | 0.01 | | 0.00 |
| 89 | F-12: Socioemotional Unresponsiveness | <-- | F-07: Motor Overflow (Excessive Impulsive Activity) | 0.01 | 0.99 | | | 0.00 | 0.00 | | 0.01 | | | | |
| 90 | F-14: Body/Head Movements (Repetitive Rocking/Turning) | <-- | F-06: Social Atypicalities (Awkward, Odd Responses) | 0.01 | 0.98 | | | 0.00 | 0.00 | 0.00 | 0.01 | | | | |
| 91 | F-17: Sensory & Object Preoccupation | --> | F-06: Social Atypicalities (Awkward, Odd Responses) | 0.01 | 0.98 | | | 0.00 | 0.00 | 0.01 | 0.01 | | | | |
| 92 | F-12: Socioemotional Unresponsiveness | <-o | NVIQ | 0.01 | 0.99 | | | | | | | | 0.01 | | 0.00 |
| 93 | F-03: Hand/Body Movements (Recurring Mannerisms, Stereotypies) | --> | F-04: Self-Injurious Behaviors | 0.01 | 0.99 | | | 0.00 | | 0.01 | | | | | |
| 94 | F-14: Body/Head Movements (Repetitive Rocking/Turning) | <-- | F-23: Obsessive Compulsive Behaviors | 0.01 | 0.98 | | | | 0.00 | 0.01 | 0.01 | | | 0.00 | |
| 95 | F-14: Body/Head Movements (Repetitive Rocking/Turning) | <-o | NVIQ | 0.01 | 0.99 | | | | | | | | 0.01 | | |
| 96 | F-16: Self-Confidence (SCI) | <-- | F-17: Sensory & Object Preoccupation | 0.01 | 0.99 | | | | 0.00 | | 0.01 | | | | |
| 97 | F-12: Socioemotional Unresponsiveness | <-- | F-15: Socioemotional Awareness (Responsive/Expressive) | 0.01 | 0.98 | | | 0.00 | 0.00 | 0.01 | 0.01 | | | | |
| 98 | F-14: Body/Head Movements (Repetitive Rocking/Turning) | --> | F-02: Isolated (Alone Preferred) | 0.01 | 0.99 | | | 0.00 | 0.00 | 0.01 | 0.00 | | | | |
| 99 | F-17: Sensory & Object Preoccupation | --> | F-08: Repetitive Speech (Perseverative Vocal Overflow) | 0.01 | 0.99 | | | 0.00 | | 0.01 | 0.00 | | | | |
| 100 | F-01: Oppositional (Outburst Behaviors) | <-- | F-02: Isolated (Alone Preferred) | 0.01 | 0.99 | | | | 0.00 | 0.00 | 0.01 | | | | |
| 101 | F-03: Hand/Body Movements (Recurring Mannerisms, Stereotypies) | --> | F-06: Social Atypicalities (Awkward, Odd Responses) | 0.01 | 0.99 | | | | | 0.01 | 0.00 | | | | |
| 102 | F-15: Socioemotional Awareness (Responsive/Expressive) | --> | F-04: Self-Injurious Behaviors | 0.01 | 0.99 | | | 0.00 | | 0.01 | 0.00 | | | | |
| 103 | AGE | o-> | F-08: Repetitive Speech (Perseverative Vocal Overflow) | 0.01 | 0.99 | | | | | 0.00 | | 0.01 | | | |
| 104 | F-01: Oppositional (Outburst Behaviors) | --> | F-03: Hand/Body Movements (Recurring Mannerisms, Stereotypies) | 0.01 | 0.99 | | | | | 0.01 | 0.00 | | | | |
| 105 | F-04: Self-Injurious Behaviors | <-- | F-05: Inflexible (Insistent Behaviors) | 0.01 | 0.99 | | | | | 0.00 | 0.01 | | | | |
| 106 | F-23: Obsessive Compulsive Behaviors | <-- | F-06: Social Atypicalities (Awkward, Odd Responses) | 0.01 | 0.99 | | | | 0.01 | | 0.01 | | | | |
| 107 | F-02: Isolated (Alone Preferred) | <-o | NVIQ | 0.01 | 0.99 | | 0.00 | | | | | | 0.01 | | 0.00 |
| 108 | F-05: Inflexible (Insistent Behaviors) | <-o | NVIQ | 0.01 | 1.00 | | | | | | | | 0.01 | | |
| 109 | F-12: Socioemotional Unresponsiveness | --> | F-17: Sensory & Object Preoccupation | 0.01 | 0.99 | | | 0.00 | 0.00 | 0.01 | 0.00 | | | | |
| 110 | F-16: Self-Confidence (SCI) | <-- | F-23: Obsessive Compulsive Behaviors | 0.01 | 0.99 | | | | 0.00 | 0.00 | 0.01 | | | | |
| 111 | F-12: Socioemotional Unresponsiveness | --> | F-04: Self-Injurious Behaviors | 0.00 | 1.00 | | | | | 0.00 | | | | | |
| 112 | F-12: Socioemotional Unresponsiveness | <-- | F-03: Hand/Body Movements (Recurring Mannerisms, Stereotypies) | 0.00 | 0.99 | | | 0.00 | 0.00 | | 0.00 | | 0.00 | | |
| 113 | F-17: Sensory & Object Preoccupation | --> | F-01: Oppositional (Outburst Behaviors) | 0.00 | 1.00 | | | | | 0.00 | | | | | |
| 114 | AGE | o-> | F-03: Hand/Body Movements (Recurring Mannerisms, Stereotypies) | 0.00 | 1.00 | | | | | 0.00 | | 0.00 | | | |
| 115 | F-10: Staring (into Space; Preoccupied) | <-- | F-16: Self-Confidence (SCI) | 0.00 | 1.00 | | | | | | 0.00 | | | | |
| 116 | F-10: Staring (into Space; Preoccupied) | <-- | F-23: Obsessive Compulsive Behaviors | 0.00 | 1.00 | | | | | | 0.00 | | | | |
| 117 | AGE | o-> | F-01: Oppositional (Outburst Behaviors) | 0.00 | 1.00 | | | | | | | 0.00 | | | |
| 118 | F-01: Oppositional (Outburst Behaviors) | --> | F-23: Obsessive Compulsive Behaviors | 0.00 | 1.00 | | | | | 0.00 | 0.00 | | | | |
| 119 | F-01: Oppositional (Outburst Behaviors) | <-o | NVIQ | 0.00 | 1.00 | | | | | | | | 0.00 | | |
| 120 | F-04: Self-Injurious Behaviors | <-- | F-06: Social Atypicalities (Awkward, Odd Responses) | 0.00 | 1.00 | | | | | | 0.00 | | | | |
| 121 | F-05: Inflexible (Insistent Behaviors) | <-- | F-06: Social Atypicalities (Awkward, Odd Responses) | 0.00 | 1.00 | | | | | | 0.00 | | | | |
| 122 | F-12: Socioemotional Unresponsiveness | --> | F-08: Repetitive Speech (Perseverative Vocal Overflow) | 0.00 | 1.00 | | | | | 0.00 | 0.00 | | | | |
| 123 | F-14: Body/Head Movements (Repetitive Rocking/Turning) | --> | F-16: Self-Confidence (SCI) | 0.00 | 1.00 | | | 0.00 | | 0.00 | 0.00 | | | | |
| 124 | AGE | o-> | F-23: Obsessive Compulsive Behaviors | 0.00 | 1.00 | | | | | | | 0.00 | | | |
| 125 | F-01: Oppositional (Outburst Behaviors) | <-o | VIQ | 0.00 | 1.00 | | | | | | | | 0.00 | | |
| 126 | F-06: Social Atypicalities (Awkward, Odd Responses) | --> | F-07: Motor Overflow (Excessive Impulsive Activity) | 0.00 | 1.00 | | | | | 0.00 | | | | | |
| 127 | F-06: Social Atypicalities (Awkward, Odd Responses) | <-o | VIQ | 0.00 | 1.00 | | | | | | | | 0.00 | | |
| 128 | F-10: Staring (into Space; Preoccupied) | --> | F-12: Socioemotional Unresponsiveness | 0.00 | 1.00 | | | | | 0.00 | | | | | |
| 129 | F-10: Staring (into Space; Preoccupied) | --> | F-15: Socioemotional Awareness (Responsive/Expressive) | 0.00 | 1.00 | | | | | 0.00 | 0.00 | | | | |
| 130 | F-10: Staring (into Space; Preoccupied) | <-- | F-04: Self-Injurious Behaviors | 0.00 | 1.00 | | | | | | 0.00 | | | | |
| 131 | F-10: Staring (into Space; Preoccupied) | <-- | F-07: Motor Overflow (Excessive Impulsive Activity) | 0.00 | 1.00 | | | | | | 0.00 | | | | |
| 132 | F-10: Staring (into Space; Preoccupied) | <-- | F-05: Inflexible (Insistent Behaviors) | 0.00 | 1.00 | | | | 0.00 | | | | | | |
| 133 | F-14: Body/Head Movements (Repetitive Rocking/Turning) | <-- | F-01: Oppositional (Outburst Behaviors) | 0.00 | 1.00 | | | | | | 0.00 | | | | |
| 134 | F-14: Body/Head Movements (Repetitive Rocking/Turning) | <-- | F-15: Socioemotional Awareness (Responsive/Expressive) | 0.00 | 1.00 | | | | 0.00 | | | | | | |
| 135 | F-15: Socioemotional Awareness (Responsive/Expressive) | <-- | F-08: Repetitive Speech (Perseverative Vocal Overflow) | 0.00 | 1.00 | | | | | | 0.00 | | | | |
| 136 | F-15: Socioemotional Awareness (Responsive/Expressive) | --> | F-23: Obsessive Compulsive Behaviors | 0.00 | 1.00 | | | 0.00 | | | | | | | |
| 137 | F-16: Self-Confidence (SCI) | <-- | F-01: Oppositional (Outburst Behaviors) | 0.00 | 1.00 | | | | | | 0.00 | | | | |
| 138 | F-16: Self-Confidence (SCI) | <-- | F-03: Hand/Body Movements (Recurring Mannerisms, Stereotypies) | 0.00 | 1.00 | | | | | | 0.00 | | | | |
| 139 | F-16: Self-Confidence (SCI) | <-- | F-05: Inflexible (Insistent Behaviors) | 0.00 | 1.00 | | | | | | 0.00 | | | | |
| 140 | F-23: Obsessive Compulsive Behaviors | <-- | F-03: Hand/Body Movements (Recurring Mannerisms, Stereotypies) | 0.00 | 1.00 | | | | | | 0.00 | | | | |

### B. MATCHED MALES

| | Node 1 | Interaction | Node 2 | Ensemble | No Edge | --> | <-- | --> | <-- | --> | <-- | o-> | <-o | o-o | --- |
|---|---|---|---|---|---|---|---|---|---|---|---|---|---|---|---|
| 1 | F-17: Sensory & Object Preoccupation | --> | F-23: Obsessive Compulsive Behaviors | 0.97 | 0.00 | | | 0.02 | 0.00 | 0.97 | 0.01 | | 0.00 | | |
| 2 | F-01: Oppositional (Outburst Behaviors) | <-- | F-07: Motor Overflow (Excessive Impulsive Activity) | 0.90 | 0.00 | | | 0.00 | 0.08 | 0.02 | 0.90 | | | | |
| 3 | F-23: Obsessive Compulsive Behaviors | --> | F-08: Repetitive Speech (Perseverative Vocal Overflow) | 0.89 | 0.00 | | | 0.08 | 0.00 | 0.89 | 0.02 | 0.00 | | | 0.01 |
| 4 | F-16: Self-Confidence (SCI) | <-o | NVIQ | 0.86 | 0.08 | | 0.03 | | 0.01 | | 0.00 | | 0.86 | | 0.02 |
| 5 | F-01: Oppositional (Outburst Behaviors) | --> | F-04: Self-Injurious Behaviors | 0.81 | 0.03 | | | 0.08 | 0.05 | 0.81 | 0.02 | 0.00 | | | 0.01 |
| 6 | F-14: Body/Head Movements (Repetitive Rocking/Turning) | --> | F-04: Self-Injurious Behaviors | 0.67 | 0.08 | 0.00 | | 0.67 | 0.00 | 0.22 | 0.00 | 0.02 | | | 0.00 |
| 7 | F-12: Socioemotional Unresponsiveness | <-- | F-02: Isolated (Alone Preferred) | 0.62 | 0.00 | | 0.00 | 0.01 | 0.11 | 0.09 | 0.62 | | 0.05 | | 0.12 |
| 8 | F-12: Socioemotional Unresponsiveness | <-o | VIQ | 0.62 | 0.09 | | 0.01 | | | | | | 0.62 | | 0.28 |
| 9 | F-03: Hand/Body Movements (Recurring Mannerisms, Stereotypies) | --> | F-08: Repetitive Speech (Perseverative Vocal Overflow) | 0.59 | 0.00 | 0.00 | | 0.59 | | 0.38 | 0.01 | 0.01 | | | 0.01 |
| 10 | F-07: Motor Overflow (Excessive Impulsive Activity) | <-- | F-08: Repetitive Speech (Perseverative Vocal Overflow) | 0.59 | 0.06 | | | 0.01 | 0.20 | 0.15 | 0.59 | | | | 0.00 |
| 11 | F-14: Body/Head Movements (Repetitive Rocking/Turning) | <-- | F-03: Hand/Body Movements (Recurring Mannerisms, Stereotypies) | 0.57 | 0.00 | 0.02 | | 0.00 | 0.57 | 0.03 | 0.33 | 0.01 | 0.04 | 0.00 | 0.01 |
| 12 | F-04: Self-Injurious Behaviors | <-o | NVIQ | 0.55 | 0.33 | | 0.10 | | 0.01 | | 0.01 | | 0.55 | | |
| 13 | F-14: Body/Head Movements (Repetitive Rocking/Turning) | --> | F-07: Motor Overflow (Excessive Impulsive Activity) | 0.54 | 0.32 | | | 0.13 | 0.00 | 0.54 | | 0.01 | | | 0.00 |
| 14 | F-14: Body/Head Movements (Repetitive Rocking/Turning) | <-- | F-05: Inflexible (Insistent Behaviors) | 0.53 | 0.22 | 0.00 | | 0.13 | 0.02 | 0.05 | 0.53 | 0.01 | 0.00 | | 0.04 |
| 15 | AGE | <-> | F-17: Sensory & Object Preoccupation | 0.52 | 0.00 | 0.01 | | 0.01 | | 0.06 | | 0.40 | | | 0.52 |
| 16 | F-23: Obsessive Compulsive Behaviors | --> | F-05: Inflexible (Insistent Behaviors) | 0.51 | 0.00 | | | 0.44 | | 0.51 | 0.04 | 0.00 | 0.00 | | 0.01 |
| 17 | F-17: Sensory & Object Preoccupation | <-o | NVIQ | 0.50 | 0.47 | | 0.02 | | 0.00 | | 0.01 | | 0.50 | | 0.01 |
| 18 | F-03: Hand/Body Movements (Recurring Mannerisms, Stereotypies) | <-o | NVIQ | 0.48 | 0.29 | | 0.20 | | 0.01 | | 0.01 | | 0.48 | | 0.00 |
| 19 | F-02: Isolated (Alone Preferred) | <-- | F-06: Social Atypicalities (Awkward, Odd Responses) | 0.47 | 0.00 | 0.00 | | 0.12 | 0.03 | 0.33 | 0.47 | 0.03 | | | 0.02 |
| 20 | F-12: Socioemotional Unresponsiveness | --> | F-16: Self-Confidence (SCI) | 0.44 | 0.45 | | | 0.06 | 0.00 | 0.44 | 0.03 | | | | 0.02 |
| 21 | F-15: Socioemotional Awareness (Responsive/Expressive) | <-o | NVIQ | 0.43 | 0.54 | | 0.02 | | 0.00 | | 0.01 | | 0.43 | | 0.00 |
| 22 | F-16: Self-Confidence (SCI) | <-- | F-02: Isolated (Alone Preferred) | 0.43 | 0.00 | | 0.01 | | 0.36 | 0.01 | 0.43 | 0.00 | 0.00 | | 0.19 |
| 23 | F-01: Oppositional (Outburst Behaviors) | <-- | F-05: Inflexible (Insistent Behaviors) | 0.42 | 0.04 | | | 0.01 | 0.42 | 0.17 | 0.33 | | 0.00 | | 0.02 |
| 24 | AGE | o-> | F-07: Motor Overflow (Excessive Impulsive Activity) | 0.40 | 0.00 | 0.01 | | 0.28 | | 0.28 | | 0.40 | | | 0.04 |
| 25 | F-15: Socioemotional Awareness (Responsive/Expressive) | --> | F-06: Social Atypicalities (Awkward, Odd Responses) | 0.40 | 0.00 | | | 0.01 | 0.03 | 0.40 | 0.18 | 0.02 | | 0.00 | 0.36 |
| 26 | F-17: Sensory & Object Preoccupation | --> | F-03: Hand/Body Movements (Recurring Mannerisms, Stereotypies) | 0.39 | 0.12 | | 0.00 | 0.04 | 0.02 | 0.39 | 0.34 | 0.00 | 0.04 | | 0.03 |
| 27 | F-10: Staring (into Space; Preoccupied) | <-- | F-08: Repetitive Speech (Perseverative Vocal Overflow) | 0.38 | 0.29 | 0.01 | | 0.11 | 0.08 | 0.06 | 0.38 | 0.04 | 0.00 | | 0.04 |
| 28 | AGE | o-> | F-12: Socioemotional Unresponsiveness | 0.38 | 0.00 | 0.03 | | 0.08 | | 0.15 | | 0.38 | | | 0.37 |
| 29 | F-10: Staring (into Space; Preoccupied) | --> | F-02: Isolated (Alone Preferred) | 0.36 | 0.00 | 0.03 | | 0.21 | 0.05 | 0.36 | 0.16 | 0.04 | 0.01 | 0.01 | 0.14 |
| 30 | AGE | <-> | F-06: Social Atypicalities (Awkward, Odd Responses) | 0.35 | 0.00 | 0.09 | | 0.09 | | 0.15 | | 0.31 | | | 0.35 |
| 31 | F-06: Social Atypicalities (Awkward, Odd Responses) | <-- | F-08: Repetitive Speech (Perseverative Vocal Overflow) | 0.34 | 0.20 | | | 0.08 | 0.35 | 0.08 | 0.15 | | | | 0.16 |
| 32 | F-12: Socioemotional Unresponsiveness | --> | F-05: Inflexible (Insistent Behaviors) | 0.31 | 0.50 | | | 0.09 | 0.03 | 0.31 | 0.02 | | | | 0.07 |
| 33 | F-12: Socioemotional Unresponsiveness | --> | F-01: Oppositional (Outburst Behaviors) | 0.29 | 0.41 | | | 0.30 | 0.01 | 0.24 | 0.03 | | | | 0.01 |
| 34 | F-14: Body/Head Movements (Repetitive Rocking/Turning) | <-- | F-17: Sensory & Object Preoccupation | 0.29 | 0.44 | | | 0.00 | 0.29 | 0.13 | 0.06 | 0.02 | | | 0.05 |
| 35 | F-10: Staring (into Space; Preoccupied) | --> | F-12: Socioemotional Unresponsiveness | 0.29 | 0.37 | 0.02 | | 0.29 | 0.00 | 0.16 | 0.01 | 0.01 | | | 0.14 |
| 36 | F-10: Staring (into Space; Preoccupied) | <-- | F-03: Hand/Body Movements (Recurring Mannerisms, Stereotypies) | 0.27 | 0.35 | 0.00 | 0.00 | 0.02 | 0.27 | 0.18 | 0.12 | 0.02 | | | 0.04 |
| 37 | F-10: Staring (into Space; Preoccupied) | <-- | F-17: Sensory & Object Preoccupation | 0.27 | 0.64 | | | 0.02 | 0.27 | 0.03 | 0.03 | 0.01 | | | 0.01 |
| 38 | F-10: Staring (into Space; Preoccupied) | --> | F-06: Social Atypicalities (Awkward, Odd Responses) | 0.27 | 0.23 | 0.00 | | 0.27 | 0.01 | 0.17 | 0.03 | 0.04 | | | 0.26 |
| 39 | F-17: Sensory & Object Preoccupation | <-o | VIQ | 0.27 | 0.53 | | 0.00 | | 0.01 | | 0.06 | | 0.27 | | 0.13 |
| 40 | F-12: Socioemotional Unresponsiveness | --> | F-14: Body/Head Movements (Repetitive Rocking/Turning) | 0.26 | 0.53 | | 0.00 | 0.08 | 0.10 | 0.26 | 0.01 | | 0.00 | | 0.01 |
| 41 | F-15: Socioemotional Awareness (Responsive/Expressive) | <-o | VIQ | 0.24 | 0.52 | | 0.03 | | 0.00 | | 0.05 | | 0.24 | | 0.17 |
| 42 | F-02: Isolated (Alone Preferred) | --> | F-05: Inflexible (Insistent Behaviors) | 0.24 | 0.42 | 0.00 | | 0.24 | 0.09 | 0.16 | 0.06 | 0.01 | 0.00 | | 0.02 |
| 43 | F-10: Staring (into Space; Preoccupied) | <-- | F-14: Body/Head Movements (Repetitive Rocking/Turning) | 0.21 | 0.59 | 0.00 | 0.01 | 0.04 | 0.09 | 0.01 | 0.22 | 0.01 | 0.01 | | 0.02 |
| 44 | F-02: Isolated (Alone Preferred) | <-- | F-03: Hand/Body Movements (Recurring Mannerisms, Stereotypies) | 0.21 | 0.67 | 0.00 | | 0.02 | 0.21 | 0.06 | 0.02 | 0.00 | | | 0.02 |
| 45 | F-15: Socioemotional Awareness (Responsive/Expressive) | --> | F-16: Self-Confidence (SCI) | 0.20 | 0.46 | 0.00 | | 0.11 | 0.00 | 0.21 | 0.14 | 0.06 | 0.00 | | 0.02 |
| 46 | F-04: Self-Injurious Behaviors | <-o | VIQ | 0.18 | 0.72 | | 0.01 | | 0.06 | | 0.03 | | 0.18 | | |
| 47 | F-12: Socioemotional Unresponsiveness | <-> | F-15: Socioemotional Awareness (Responsive/Expressive) | 0.16 | 0.50 | | | 0.02 | 0.11 | 0.07 | 0.11 | | 0.03 | | 0.16 |
| 48 | F-03: Hand/Body Movements (Recurring Mannerisms, Stereotypies) | <-o | VIQ | 0.16 | 0.70 | | 0.05 | | 0.02 | | 0.07 | | 0.16 | | |
| 49 | F-14: Body/Head Movements (Repetitive Rocking/Turning) | <-o | NVIQ | 0.16 | 0.82 | | 0.02 | | 0.00 | | 0.00 | | 0.16 | | 0.00 |
| 50 | F-07: Motor Overflow (Excessive Impulsive Activity) | <-o | NVIQ | 0.15 | 0.83 | | 0.01 | | 0.00 | | 0.00 | | 0.15 | | 0.00 |
| 51 | F-17: Sensory & Object Preoccupation | --> | F-06: Social Atypicalities (Awkward, Odd Responses) | 0.12 | 0.85 | | | 0.12 | 0.01 | 0.01 | 0.01 | | | | |
| 52 | AGE | <-> | F-15: Socioemotional Awareness (Responsive/Expressive) | 0.12 | 0.69 | 0.08 | | 0.04 | | 0.05 | | 0.03 | | | 0.12 |
| 53 | F-04: Self-Injurious Behaviors | <-- | F-05: Inflexible (Insistent Behaviors) | 0.12 | 0.81 | | 0.00 | 0.00 | 0.12 | 0.01 | 0.04 | | | | 0.02 |
| 54 | F-01: Oppositional (Outburst Behaviors) | --> | F-06: Social Atypicalities (Awkward, Odd Responses) | 0.11 | 0.71 | | | 0.02 | 0.09 | 0.11 | 0.06 | | | | 0.01 |
| 55 | F-03: Hand/Body Movements (Recurring Mannerisms, Stereotypies) | --> | F-07: Motor Overflow (Excessive Impulsive Activity) | 0.08 | 0.87 | | | 0.08 | | 0.05 | 0.00 | 0.00 | | | |
| 56 | F-02: Isolated (Alone Preferred) | <-- | F-08: Repetitive Speech (Perseverative Vocal Overflow) | 0.08 | 0.89 | | | | 0.08 | 0.01 | 0.02 | | | | 0.01 |
| 57 | F-04: Self-Injurious Behaviors | --> | F-07: Motor Overflow (Excessive Impulsive Activity) | 0.07 | 0.92 | | | 0.00 | 0.01 | 0.07 | | 0.00 | | | 0.00 |
| 58 | F-15: Socioemotional Awareness (Responsive/Expressive) | <-- | F-17: Sensory & Object Preoccupation | 0.07 | 0.88 | | | | 0.04 | 0.01 | 0.07 | 0.00 | | 0.00 | 0.00 |
| 59 | F-03: Hand/Body Movements (Recurring Mannerisms, Stereotypies) | --> | F-04: Self-Injurious Behaviors | 0.07 | 0.92 | 0.00 | | 0.07 | | 0.00 | 0.00 | 0.01 | 0.00 | | |
| 60 | F-05: Inflexible (Insistent Behaviors) | <-o | VIQ | 0.06 | 0.92 | | 0.00 | | 0.00 | | 0.02 | | 0.06 | | |
| 61 | F-06: Social Atypicalities (Awkward, Odd Responses) | <-o | NVIQ | 0.04 | 0.96 | | | | | | 0.00 | | 0.04 | | |
| 62 | F-16: Self-Confidence (SCI) | <-- | F-08: Repetitive Speech (Perseverative Vocal Overflow) | 0.04 | 0.93 | | | | 0.04 | | 0.04 | | | | |
| 63 | AGE | --> | F-02: Isolated (Alone Preferred) | 0.03 | 0.93 | 0.00 | | 0.01 | | 0.03 | | 0.02 | | | 0.00 |
| 64 | F-14: Body/Head Movements (Repetitive Rocking/Turning) | <-- | F-23: Obsessive Compulsive Behaviors | 0.03 | 0.92 | 0.00 | | 0.01 | 0.02 | 0.02 | 0.03 | | 0.00 | | 0.00 |
| 65 | F-14: Body/Head Movements (Repetitive Rocking/Turning) | <-> | VIQ | 0.03 | 0.92 | | 0.02 | | 0.00 | | 0.01 | | 0.02 | | 0.03 |
| 66 | F-17: Sensory & Object Preoccupation | --> | F-04: Self-Injurious Behaviors | 0.03 | 0.97 | | | 0.03 | | 0.00 | | | | | |
| 67 | F-05: Inflexible (Insistent Behaviors) | <-- | F-06: Social Atypicalities (Awkward, Odd Responses) | 0.03 | 0.95 | | | | 0.02 | | 0.03 | | | | |
| 68 | AGE | o-> | F-16: Self-Confidence (SCI) | 0.02 | 0.94 | 0.00 | | 0.02 | | 0.02 | | 0.02 | | | |

| | | | | | | | | | | | | | | | |
|---|---|---|---|---|---|---|---|---|---|---|---|---|---|---|---|
| 69 | F-08: Repetitive Speech (Perseverative Vocal Overflow) | <-o | VIQ | 0.02 | 0.96 | | 0.00 | | 0.01 | | 0.01 | | 0.02 | | |
| 70 | AGE | o-> | F-05: Inflexible (Insistent Behaviors) | 0.02 | 0.96 | | | 0.01 | | 0.00 | | 0.02 | | | |
| 71 | F-05: Inflexible (Insistent Behaviors) | <-- | F-08: Repetitive Speech (Perseverative Vocal Overflow) | 0.02 | 0.97 | | | | 0.01 | 0.00 | 0.02 | 0.00 | | | |
| 72 | F-15: Socioemotional Awareness (Responsive/Expressive) | <-- | F-05: Inflexible (Insistent Behaviors) | 0.02 | 0.97 | | | 0.00 | 0.01 | 0.00 | 0.02 | | | | |
| 73 | F-08: Repetitive Speech (Perseverative Vocal Overflow) | <-> | NVIQ | 0.02 | 0.97 | | 0.00 | | | | | | 0.02 | | 0.02 |
| 74 | F-23: Obsessive Compulsive Behaviors | --> | F-02: Isolated (Alone Preferred) | 0.02 | 0.95 | | | 0.02 | 0.01 | 0.01 | 0.01 | | 0.00 | | |
| 75 | F-12: Socioemotional Unresponsiveness | <-o | NVIQ | 0.01 | 0.98 | | | | | | | | 0.01 | | 0.01 |
| 76 | F-16: Self-Confidence (SCI) | --> | F-06: Social Atypicalities (Awkward, Odd Responses) | 0.01 | 0.98 | | | | | 0.01 | 0.00 | | | | 0.01 |
| 77 | AGE | o-> | F-08: Repetitive Speech (Perseverative Vocal Overflow) | 0.01 | 0.98 | | | 0.01 | | 0.00 | | 0.01 | | | 0.00 |
| 78 | F-14: Body/Head Movements (Repetitive Rocking/Turning) | --> | F-02: Isolated (Alone Preferred) | 0.01 | 0.98 | 0.00 | | 0.01 | | 0.01 | 0.00 | | | | |
| 79 | F-16: Self-Confidence (SCI) | <-o | VIQ | 0.01 | 0.98 | | 0.00 | | 0.00 | | 0.00 | | 0.01 | | 0.00 |
| 80 | AGE | --> | F-01: Oppositional (Outburst Behaviors) | 0.01 | 0.99 | 0.01 | | 0.01 | | | | | | | |
| 81 | F-04: Self-Injurious Behaviors | <-- | F-08: Repetitive Speech (Perseverative Vocal Overflow) | 0.01 | 0.99 | | | | 0.01 | 0.00 | 0.00 | | | | |
| 82 | F-02: Isolated (Alone Preferred) | <-o | VIQ | 0.01 | 1.00 | | | | | | | | 0.01 | | |
| 83 | F-10: Staring (into Space; Preoccupied) | <-- | F-15: Socioemotional Awareness (Responsive/Expressive) | 0.01 | 0.99 | | | | 0.01 | | 0.00 | | | | |
| 84 | F-12: Socioemotional Unresponsiveness | --> | F-23: Obsessive Compulsive Behaviors | 0.01 | 0.99 | | | 0.00 | 0.00 | 0.01 | | | | | |
| 85 | F-16: Self-Confidence (SCI) | <-- | F-07: Motor Overflow (Excessive Impulsive Activity) | 0.01 | 0.99 | | | | 0.00 | | 0.01 | | | | |
| 86 | F-15: Socioemotional Awareness (Responsive/Expressive) | --> | F-23: Obsessive Compulsive Behaviors | 0.00 | 1.00 | | | | 0.00 | 0.00 | | | | | |
| 87 | F-23: Obsessive Compulsive Behaviors | <-o | VIQ | 0.00 | 1.00 | | | | | | 0.00 | | 0.00 | | |
| 88 | F-04: Self-Injurious Behaviors | --> | F-06: Social Atypicalities (Awkward, Odd Responses) | 0.00 | 0.99 | | | | 0.00 | 0.00 | 0.00 | | | | |
| 89 | F-14: Body/Head Movements (Repetitive Rocking/Turning) | --> | F-15: Socioemotional Awareness (Responsive/Expressive) | 0.00 | 1.00 | | | | | 0.00 | | | | | |
| 90 | F-23: Obsessive Compulsive Behaviors | --> | F-04: Self-Injurious Behaviors | 0.00 | 1.00 | | | 0.00 | | 0.00 | 0.00 | | | | |
| 91 | AGE | <-> | F-10: Staring (into Space; Preoccupied) | 0.00 | 0.99 | 0.00 | | 0.00 | | 0.00 | | | | | 0.00 |
| 92 | F-05: Inflexible (Insistent Behaviors) | <-o | NVIQ | 0.00 | 1.00 | | | | | | | | 0.00 | | |
| 93 | F-07: Motor Overflow (Excessive Impulsive Activity) | <-o | VIQ | 0.00 | 1.00 | | | | | | | | 0.00 | | |
| 94 | F-15: Socioemotional Awareness (Responsive/Expressive) | --> | F-07: Motor Overflow (Excessive Impulsive Activity) | 0.00 | 0.99 | | | 0.00 | | 0.00 | 0.00 | | | | |
| 95 | F-16: Self-Confidence (SCI) | --> | F-01: Oppositional (Outburst Behaviors) | 0.00 | 1.00 | | | | | 0.00 | | | | | |
| 96 | F-17: Sensory & Object Preoccupation | <-- | F-02: Isolated (Alone Preferred) | 0.00 | 1.00 | | | | 0.00 | | | | | | 0.00 |
| 97 | AGE | o-> | F-14: Body/Head Movements (Repetitive Rocking/Turning) | 0.00 | 1.00 | | | | | | | 0.00 | | | |
| 98 | AGE | o-> | F-03: Hand/Body Movements (Recurring Mannerisms, Stereotypies) | 0.00 | 1.00 | | | | | | | 0.00 | | | |
| 99 | F-01: Oppositional (Outburst Behaviors) | <-- | F-02: Isolated (Alone Preferred) | 0.00 | 1.00 | | | | 0.00 | | | | 0.00 | | |
| 100 | F-10: Staring (into Space; Preoccupied) | <-- | F-01: Oppositional (Outburst Behaviors) | 0.00 | 1.00 | | | 0.00 | 0.00 | | 0.00 | | | | |
| 101 | F-10: Staring (into Space; Preoccupied) | <-- | F-07: Motor Overflow (Excessive Impulsive Activity) | 0.00 | 1.00 | | | | 0.00 | | | | | | |
| 102 | F-10: Staring (into Space; Preoccupied) | <-o | NVIQ | 0.00 | 1.00 | | | | | | | | 0.00 | | |
| 103 | F-15: Socioemotional Awareness (Responsive/Expressive) | --> | F-01: Oppositional (Outburst Behaviors) | 0.00 | 1.00 | | | | | 0.00 | 0.00 | | | | |
| 104 | F-15: Socioemotional Awareness (Responsive/Expressive) | --> | F-03: Hand/Body Movements (Recurring Mannerisms, Stereotypies) | 0.00 | 1.00 | | | | | 0.00 | 0.00 | | | | |
| 105 | F-16: Self-Confidence (SCI) | --> | F-04: Self-Injurious Behaviors | 0.00 | 1.00 | | | | | 0.00 | | | | | |
| 106 | F-17: Sensory & Object Preoccupation | --> | F-07: Motor Overflow (Excessive Impulsive Activity) | 0.00 | 1.00 | | | 0.00 | | | | | | | |
| 107 | F-17: Sensory & Object Preoccupation | --> | F-08: Repetitive Speech (Perseverative Vocal Overflow) | 0.00 | 1.00 | | | 0.00 | | | | | | | |

### C. ALL MALES

| | Node 1 | Interaction | Node 2 | Ensemble | No Edge | --> | <-- | --> | <-- | --> | <-- | o-> | <-o | o-o | --- |
|---|---|---|---|---|---|---|---|---|---|---|---|---|---|---|---|
| 1 | F-16: Self-Confidence (SCI) | <-o | NVIQ | 0.94 | 0.03 | | 0.04 | | | | | | 0.94 | | |
| 2 | F-01: Oppositional (Outburst Behaviors) | <-- | F-07: Motor Overflow (Excessive Impulsive Activity) | 0.93 | 0.00 | | | | 0.05 | 0.02 | 0.93 | | | | |
| 3 | F-17: Sensory & Object Preoccupation | --> | F-23: Obsessive Compulsive Behaviors | 0.92 | 0.00 | | | 0.04 | | 0.92 | 0.02 | | 0.03 | | 0.00 |
| 4 | F-17: Sensory & Object Preoccupation | <-o | NVIQ | 0.88 | 0.11 | | 0.02 | | | | | | 0.88 | | |
| 5 | F-23: Obsessive Compulsive Behaviors | --> | F-08: Repetitive Speech (Perseverative Vocal Overflow) | 0.87 | 0.00 | | | 0.07 | | 0.87 | 0.04 | 0.03 | | | 0.00 |
| 6 | F-12: Socioemotional Unresponsiveness | <-o | VIQ | 0.81 | 0.01 | | 0.00 | | | | 0.00 | | 0.81 | | 0.18 |
| 7 | F-01: Oppositional (Outburst Behaviors) | --> | F-04: Self-Injurious Behaviors | 0.80 | 0.02 | | | 0.06 | 0.09 | 0.80 | 0.02 | | | | 0.00 |
| 8 | F-12: Socioemotional Unresponsiveness | <-- | F-02: Isolated (Alone Preferred) | 0.72 | 0.00 | | | 0.00 | 0.11 | 0.03 | 0.72 | | 0.04 | | 0.10 |
| 9 | AGE | <-> | F-17: Sensory & Object Preoccupation | 0.70 | 0.00 | 0.02 | | 0.02 | | 0.05 | | 0.21 | | | 0.70 |
| 10 | F-15: Socioemotional Awareness (Responsive/Expressive) | <-o | NVIQ | 0.69 | 0.28 | | 0.02 | | | | | | 0.69 | | |
| 11 | F-14: Body/Head Movements (Repetitive Rocking/Turning) | --> | F-04: Self-Injurious Behaviors | 0.67 | 0.05 | | | 0.67 | | 0.27 | 0.00 | 0.01 | | 0.00 | 0.00 |
| 12 | F-07: Motor Overflow (Excessive Impulsive Activity) | <-- | F-08: Repetitive Speech (Perseverative Vocal Overflow) | 0.66 | 0.05 | | | 0.01 | 0.16 | 0.11 | 0.66 | | | | 0.01 |
| 13 | F-14: Body/Head Movements (Repetitive Rocking/Turning) | <-- | F-03: Hand/Body Movements (Recurring Mannerisms, Stereotypies) | 0.65 | 0.00 | 0.01 | 0.00 | 0.00 | 0.65 | 0.01 | 0.31 | 0.00 | 0.01 | | 0.00 |
| 14 | F-14: Body/Head Movements (Repetitive Rocking/Turning) | <-- | F-05: Inflexible (Insistent Behaviors) | 0.65 | 0.11 | | | 0.12 | 0.03 | 0.07 | 0.65 | 0.00 | 0.00 | | 0.02 |
| 15 | F-23: Obsessive Compulsive Behaviors | --> | F-05: Inflexible (Insistent Behaviors) | 0.61 | 0.00 | | | 0.33 | 0.00 | 0.61 | 0.03 | 0.03 | | | 0.00 |
| 16 | F-03: Hand/Body Movements (Recurring Mannerisms, Stereotypies) | --> | F-08: Repetitive Speech (Perseverative Vocal Overflow) | 0.61 | 0.00 | | | 0.61 | | 0.38 | 0.01 | | | | 0.00 |
| 17 | AGE | <-> | F-12: Socioemotional Unresponsiveness | 0.59 | 0.00 | 0.03 | | 0.06 | | 0.12 | | 0.20 | | | 0.59 |
| 18 | F-14: Body/Head Movements (Repetitive Rocking/Turning) | --> | F-07: Motor Overflow (Excessive Impulsive Activity) | 0.56 | 0.30 | | | 0.13 | 0.00 | 0.56 | | 0.01 | | | |
| 19 | F-04: Self-Injurious Behaviors | <-o | NVIQ | 0.55 | 0.34 | | 0.11 | | | | | | 0.55 | | |
| 20 | F-01: Oppositional (Outburst Behaviors) | <-- | F-05: Inflexible (Insistent Behaviors) | 0.52 | 0.02 | | | | 0.52 | 0.13 | 0.31 | | 0.00 | | 0.02 |
| 21 | F-02: Isolated (Alone Preferred) | <-- | F-06: Social Atypicalities (Awkward, Odd Responses) | 0.51 | 0.00 | 0.00 | | 0.11 | 0.11 | 0.24 | 0.51 | 0.03 | 0.00 | | 0.01 |
| 22 | F-12: Socioemotional Unresponsiveness | --> | F-05: Inflexible (Insistent Behaviors) | 0.49 | 0.25 | | 0.00 | 0.10 | 0.02 | 0.49 | 0.02 | | | | 0.13 |
| 23 | F-16: Self-Confidence (SCI) | <-- | F-02: Isolated (Alone Preferred) | 0.48 | 0.00 | | 0.00 | | 0.38 | 0.01 | 0.49 | 0.00 | 0.00 | | 0.12 |
| 24 | F-15: Socioemotional Awareness (Responsive/Expressive) | --> | F-06: Social Atypicalities (Awkward, Odd Responses) | 0.47 | 0.00 | | | 0.01 | 0.05 | 0.47 | 0.20 | 0.01 | | | 0.26 |
| 25 | F-17: Sensory & Object Preoccupation | --> | F-03: Hand/Body Movements (Recurring Mannerisms, Stereotypies) | 0.46 | 0.09 | | 0.00 | 0.03 | 0.03 | 0.46 | 0.34 | | 0.02 | | 0.04 |
| 26 | AGE | <-> | F-06: Social Atypicalities (Awkward, Odd Responses) | 0.43 | 0.00 | 0.05 | | 0.12 | | 0.22 | | 0.18 | | | 0.43 |
| 27 | AGE | --> | F-07: Motor Overflow (Excessive Impulsive Activity) | 0.38 | 0.00 | 0.00 | | 0.38 | | 0.35 | | 0.23 | | | 0.05 |
| 28 | F-12: Socioemotional Unresponsiveness | --> | F-01: Oppositional (Outburst Behaviors) | 0.37 | 0.43 | | | 0.37 | 0.01 | 0.18 | 0.00 | | | | 0.01 |
| 29 | F-12: Socioemotional Unresponsiveness | --> | F-16: Self-Confidence (SCI) | 0.37 | 0.54 | | | 0.03 | 0.00 | 0.37 | 0.05 | | | | 0.01 |
| 30 | F-03: Hand/Body Movements (Recurring Mannerisms, Stereotypies) | <-o | NVIQ | 0.37 | 0.30 | | 0.34 | | | | | | 0.37 | | |

| | | | | | | | | | | | | | | | |
|---|---|---|---|---|---|---|---|---|---|---|---|---|---|---|---|
| 31 | F-10: Staring (into Space; Preoccupied) | --> | F-12: Socioemotional Unresponsiveness | 0.30 | 0.41 | 0.02 | | 0.30 | 0.01 | 0.08 | 0.02 | 0.01 | | | 0.16 |
| 32 | F-10: Staring (into Space; Preoccupied) | --> | F-02: Isolated (Alone Preferred) | 0.30 | 0.00 | 0.05 | | 0.21 | 0.04 | 0.30 | 0.12 | 0.11 | 0.01 | 0.00 | 0.18 |
| 33 | F-15: Socioemotional Awareness (Responsive/Expressive) | --> | F-16: Self-Confidence (SCI) | 0.29 | 0.11 | 0.01 | | 0.15 | 0.01 | 0.29 | 0.17 | 0.20 | 0.00 | 0.00 | 0.07 |
| 34 | F-10: Staring (into Space; Preoccupied) | <-- | F-17: Sensory & Object Preoccupation | 0.28 | 0.58 | | | 0.03 | 0.28 | 0.04 | 0.03 | 0.02 | | | 0.02 |
| 35 | F-02: Isolated (Alone Preferred) | <-- | F-03: Hand/Body Movements (Recurring Mannerisms, Stereotypies) | 0.27 | 0.45 | | | 0.03 | 0.28 | 0.17 | 0.06 | | | | 0.02 |
| 36 | F-14: Body/Head Movements (Repetitive Rocking/Turning) | <-- | F-17: Sensory & Object Preoccupation | 0.27 | 0.52 | | | 0.00 | 0.27 | 0.09 | 0.04 | 0.02 | | | 0.05 |
| 37 | F-03: Hand/Body Movements (Recurring Mannerisms, Stereotypies) | <-o | VIQ | 0.27 | 0.67 | | 0.00 | | 0.00 | | 0.05 | | 0.27 | | 0.00 |
| 38 | F-10: Staring (into Space; Preoccupied) | <-- | F-08: Repetitive Speech (Perseverative Vocal Overflow) | 0.26 | 0.37 | 0.02 | | 0.20 | 0.03 | 0.06 | 0.26 | 0.05 | 0.00 | | 0.03 |
| 39 | F-10: Staring (into Space; Preoccupied) | <-> | F-06: Social Atypicalities (Awkward, Odd Responses) | 0.25 | 0.15 | 0.00 | | 0.24 | 0.01 | 0.20 | 0.10 | 0.05 | | | 0.25 |
| 40 | F-12: Socioemotional Unresponsiveness | --> | F-14: Body/Head Movements (Repetitive Rocking/Turning) | 0.25 | 0.44 | | | 0.21 | 0.08 | 0.25 | 0.01 | | 0.00 | | 0.02 |
| 41 | F-04: Self-Injurious Behaviors | <-o | VIQ | 0.24 | 0.68 | | 0.01 | | 0.03 | | 0.04 | | 0.24 | | 0.00 |
| 42 | F-06: Social Atypicalities (Awkward, Odd Responses) | --> | F-08: Repetitive Speech (Perseverative Vocal Overflow) | 0.23 | 0.12 | | | 0.23 | 0.23 | 0.18 | 0.10 | | | | 0.13 |
| 43 | F-10: Staring (into Space; Preoccupied) | <-- | F-03: Hand/Body Movements (Recurring Mannerisms, Stereotypies) | 0.23 | 0.19 | 0.02 | | 0.07 | 0.23 | 0.22 | 0.20 | 0.02 | | | 0.04 |
| 44 | F-14: Body/Head Movements (Repetitive Rocking/Turning) | <-o | NVIQ | 0.20 | 0.77 | | 0.03 | | | | | | 0.20 | | |
| 45 | AGE | <-> | F-15: Socioemotional Awareness (Responsive/Expressive) | 0.19 | 0.58 | 0.05 | | 0.07 | | 0.08 | | 0.03 | | | 0.19 |
| 46 | F-03: Hand/Body Movements (Recurring Mannerisms, Stereotypies) | --> | F-07: Motor Overflow (Excessive Impulsive Activity) | 0.17 | 0.76 | | | 0.17 | 0.00 | 0.07 | | 0.00 | | | |
| 47 | F-04: Self-Injurious Behaviors | <-- | F-05: Inflexible (Insistent Behaviors) | 0.16 | 0.79 | | | 0.00 | 0.16 | 0.01 | 0.03 | | 0.00 | | 0.01 |
| 48 | F-02: Isolated (Alone Preferred) | --> | F-05: Inflexible (Insistent Behaviors) | 0.16 | 0.69 | 0.00 | | 0.16 | 0.04 | 0.06 | 0.03 | 0.00 | | | 0.01 |
| 49 | F-10: Staring (into Space; Preoccupied) | <-- | F-14: Body/Head Movements (Repetitive Rocking/Turning) | 0.14 | 0.73 | 0.00 | 0.00 | 0.06 | 0.03 | 0.01 | 0.14 | 0.01 | 0.01 | | 0.01 |
| 50 | F-17: Sensory & Object Preoccupation | --> | F-06: Social Atypicalities (Awkward, Odd Responses) | 0.14 | 0.79 | | | 0.14 | 0.04 | 0.01 | 0.02 | | | | |
| 51 | F-12: Socioemotional Unresponsiveness | <-- | F-15: Socioemotional Awareness (Responsive/Expressive) | 0.13 | 0.60 | | | 0.02 | 0.13 | 0.05 | 0.09 | | 0.02 | | 0.09 |
| 52 | F-15: Socioemotional Awareness (Responsive/Expressive) | <-o | VIQ | 0.13 | 0.74 | | 0.00 | | 0.00 | | 0.01 | | 0.13 | | 0.12 |
| 53 | F-07: Motor Overflow (Excessive Impulsive Activity) | <-o | NVIQ | 0.13 | 0.85 | | 0.03 | | | | | | 0.13 | | |
| 54 | F-01: Oppositional (Outburst Behaviors) | <-- | F-06: Social Atypicalities (Awkward, Odd Responses) | 0.11 | 0.77 | | | 0.01 | 0.12 | 0.07 | 0.03 | | | | 0.01 |
| 55 | F-04: Self-Injurious Behaviors | --> | F-07: Motor Overflow (Excessive Impulsive Activity) | 0.11 | 0.87 | | | 0.00 | 0.01 | 0.11 | 0.00 | | | | |
| 56 | F-05: Inflexible (Insistent Behaviors) | <-o | VIQ | 0.09 | 0.82 | | 0.00 | | 0.01 | | 0.02 | | 0.09 | | 0.00 |
| 57 | F-06: Social Atypicalities (Awkward, Odd Responses) | <-o | NVIQ | 0.08 | 0.92 | | 0.00 | | | | | | 0.08 | | |
| 58 | F-17: Sensory & Object Preoccupation | <-> | VIQ | 0.07 | 0.88 | | 0.00 | | | | 0.00 | | 0.04 | | 0.07 |
| 59 | F-05: Inflexible (Insistent Behaviors) | <-- | F-06: Social Atypicalities (Awkward, Odd Responses) | 0.07 | 0.90 | | | | 0.07 | | 0.04 | | | | |
| 60 | F-03: Hand/Body Movements (Recurring Mannerisms, Stereotypies) | --> | F-04: Self-Injurious Behaviors | 0.06 | 0.94 | | | 0.06 | | 0.00 | 0.00 | 0.00 | | | |
| 61 | F-23: Obsessive Compulsive Behaviors | <-- | F-02: Isolated (Alone Preferred) | 0.04 | 0.88 | | | 0.03 | 0.03 | 0.02 | 0.04 | | | | |
| 62 | F-17: Sensory & Object Preoccupation | --> | F-04: Self-Injurious Behaviors | 0.04 | 0.96 | | | 0.04 | | | | | | | |
| 63 | AGE | --> | F-16: Self-Confidence (SCI) | 0.04 | 0.91 | 0.00 | | 0.03 | | 0.04 | | 0.02 | | | |
| 64 | AGE | --> | F-02: Isolated (Alone Preferred) | 0.04 | 0.90 | 0.00 | | 0.03 | | 0.04 | | 0.02 | | | 0.01 |
| 65 | F-02: Isolated (Alone Preferred) | <-- | F-08: Repetitive Speech (Perseverative Vocal Overflow) | 0.04 | 0.93 | | | 0.00 | 0.04 | 0.00 | 0.02 | | | | 0.01 |
| 66 | F-16: Self-Confidence (SCI) | <-- | F-08: Repetitive Speech (Perseverative Vocal Overflow) | 0.04 | 0.95 | | | 0.00 | 0.01 | 0.00 | 0.04 | | | | |
| 67 | F-15: Socioemotional Awareness (Responsive/Expressive) | <-- | F-17: Sensory & Object Preoccupation | 0.03 | 0.94 | | | 0.00 | 0.02 | 0.00 | 0.03 | 0.00 | 0.00 | 0.00 | |
| 68 | F-10: Staring (into Space; Preoccupied) | <-- | F-15: Socioemotional Awareness (Responsive/Expressive) | 0.03 | 0.96 | | | 0.00 | 0.03 | | 0.01 | | | | |
| 69 | F-07: Motor Overflow (Excessive Impulsive Activity) | <-o | VIQ | 0.02 | 0.97 | | | | 0.01 | | 0.00 | | 0.02 | | |
| 70 | F-14: Body/Head Movements (Repetitive Rocking/Turning) | <-- | VIQ | 0.02 | 0.96 | | 0.02 | | | | 0.00 | | 0.00 | | 0.01 |
| 71 | AGE | --> | F-01: Oppositional (Outburst Behaviors) | 0.02 | 0.98 | 0.01 | | 0.02 | | | | | | | 0.00 |
| 72 | F-15: Socioemotional Awareness (Responsive/Expressive) | <-- | F-05: Inflexible (Insistent Behaviors) | 0.02 | 0.97 | | | 0.01 | 0.01 | 0.00 | 0.02 | | | | 0.00 |
| 73 | F-05: Inflexible (Insistent Behaviors) | <-- | F-08: Repetitive Speech (Perseverative Vocal Overflow) | 0.01 | 0.98 | | | | 0.01 | | 0.01 | | | | |
| 74 | F-14: Body/Head Movements (Repetitive Rocking/Turning) | --> | F-23: Obsessive Compulsive Behaviors | 0.01 | 0.96 | | | 0.01 | 0.01 | 0.01 | 0.01 | | 0.01 | | |
| 75 | F-16: Self-Confidence (SCI) | <-- | F-07: Motor Overflow (Excessive Impulsive Activity) | 0.01 | 0.99 | | | | | | 0.01 | | | | |
| 76 | F-23: Obsessive Compulsive Behaviors | <-o | VIQ | 0.01 | 0.99 | | | | | | | | 0.01 | | |
| 77 | F-15: Socioemotional Awareness (Responsive/Expressive) | --> | F-23: Obsessive Compulsive Behaviors | 0.01 | 0.99 | | | 0.01 | | 0.00 | 0.00 | | | | |
| 78 | F-16: Self-Confidence (SCI) | --> | F-04: Self-Injurious Behaviors | 0.01 | 0.99 | | | 0.00 | 0.00 | 0.01 | 0.00 | | | | |
| 79 | F-06: Social Atypicalities (Awkward, Odd Responses) | <-o | VIQ | 0.01 | 0.99 | | | | | | 0.00 | | 0.01 | | |
| 80 | AGE | --> | F-05: Inflexible (Insistent Behaviors) | 0.01 | 0.99 | | | 0.01 | | 0.00 | | 0.00 | | | |
| 81 | F-08: Repetitive Speech (Perseverative Vocal Overflow) | <-o | VIQ | 0.01 | 0.99 | | | | | | 0.00 | | 0.01 | | |
| 82 | F-16: Self-Confidence (SCI) | --> | F-06: Social Atypicalities (Awkward, Odd Responses) | 0.01 | 0.99 | | | | 0.00 | 0.01 | 0.00 | | | | 0.00 |
| 83 | F-04: Self-Injurious Behaviors | <-- | F-06: Social Atypicalities (Awkward, Odd Responses) | 0.01 | 0.99 | | | | 0.01 | | 0.00 | | | | |
| 84 | F-10: Staring (into Space; Preoccupied) | --> | F-01: Oppositional (Outburst Behaviors) | 0.01 | 0.99 | | | 0.01 | 0.00 | 0.00 | 0.00 | | | | |
| 85 | F-17: Sensory & Object Preoccupation | <-- | F-02: Isolated (Alone Preferred) | 0.01 | 0.99 | | | | 0.00 | | 0.01 | | | | 0.00 |
| 86 | F-01: Oppositional (Outburst Behaviors) | <-- | F-02: Isolated (Alone Preferred) | 0.00 | 1.00 | | | | 0.00 | | 0.00 | | | | |
| 87 | F-14: Body/Head Movements (Repetitive Rocking/Turning) | --> | F-02: Isolated (Alone Preferred) | 0.00 | 1.00 | | | 0.00 | | 0.00 | | | | | |
| 88 | F-15: Socioemotional Awareness (Responsive/Expressive) | <-- | F-07: Motor Overflow (Excessive Impulsive Activity) | 0.00 | 1.00 | | | 0.00 | | 0.00 | 0.00 | | | | |
| 89 | F-16: Self-Confidence (SCI) | --> | F-05: Inflexible (Insistent Behaviors) | 0.00 | 1.00 | | | | | 0.00 | | | | | |
| 90 | F-01: Oppositional (Outburst Behaviors) | <-o | VIQ | 0.00 | 1.00 | | | | | | | | 0.00 | | |
| 91 | F-04: Self-Injurious Behaviors | <-- | F-08: Repetitive Speech (Perseverative Vocal Overflow) | 0.00 | 1.00 | | | | 0.00 | | 0.00 | | | | |
| 92 | F-15: Socioemotional Awareness (Responsive/Expressive) | <-- | F-03: Hand/Body Movements (Recurring Mannerisms, Stereotypies) | 0.00 | 1.00 | | | | 0.00 | | 0.00 | | | | |
| 93 | F-16: Self-Confidence (SCI) | <-- | VIQ | 0.00 | 0.99 | | 0.00 | | | | | | 0.00 | | |
| 94 | F-23: Obsessive Compulsive Behaviors | --> | F-04: Self-Injurious Behaviors | 0.00 | 1.00 | | | 0.00 | | 0.00 | | | | | |
| 95 | F-08: Repetitive Speech (Perseverative Vocal Overflow) | <-o | NVIQ | 0.00 | 1.00 | | | | | | | | 0.00 | | |
| 96 | F-12: Socioemotional Unresponsiveness | --> | F-23: Obsessive Compulsive Behaviors | 0.00 | 1.00 | | | 0.00 | | 0.00 | | | | | |
| 97 | F-16: Self-Confidence (SCI) | --> | F-01: Oppositional (Outburst Behaviors) | 0.00 | 1.00 | | | | | 0.00 | | | | | |
| 98 | F-17: Sensory & Object Preoccupation | <-- | F-05: Inflexible (Insistent Behaviors) | 0.00 | 1.00 | | | | | | 0.00 | | | | |
| 99 | F-23: Obsessive Compulsive Behaviors | <-- | F-07: Motor Overflow (Excessive Impulsive Activity) | 0.00 | 1.00 | | | 0.00 | | | 0.00 | | | | |
